# Acute exercise rewires the proteomic landscape of human immune cells

**DOI:** 10.1101/2025.01.30.25321196

**Authors:** David Walzik, Niklas Joisten, Alan J Metcalfe, Sebastian Proschinger, Alexander Schenk, Charlotte Wenzel, Alessa L. Henneberg, Martin Schneider, Silvia Calderazzo, Andreas Groll, Carsten Watzl, Christiane A. Opitz, Dominic Helm, Philipp Zimmer

## Abstract

Exercise-driven alterations of the immune system are a key mechanism in the prevention and treatment of various diseases. Here, we performed mass spectrometry-based proteomics analysis on peripheral blood mononuclear cells (PBMCs) at a depth of >6000 proteins. Comparing time– and workload-matched high-intensity interval exercise (HIIE) and moderate-intensity continuous exercise (MICE) we discover versatile changes in the proteomic makeup of PBMCs and reveal profound alterations related to effector function and immune cell activation pathways within one hour after exercise. These changes were more pronounced after HIIE compared to MICE and occurred despite identical immune cell mobilization patterns between the two exercise conditions. We further identify an immunoproteomic signature that effectively predicts cardiorespiratory fitness. This study provides a reliable data resource that expands our knowledge on how exercise modulates the immune system, and delivers biological evidence supporting the WHO 2020 guidelines, which highlight exercise intensity as a relevant factor to maintain health.

## INTRODUCTION

Physical exercise is one of the most powerful strategies to prevent and counteract acute and chronic diseases across the entire human lifespan. The health benefits of exercise are demonstrated by a vast body of evidence from clinical and observational trials,^1,2^ but the underlying biological mechanisms are poorly understood. Omics-approaches have improved our understanding of the molecular basis of exercise, however, so far these methods have only been applied comprehensively in animal models.^3,4^ In contrast, human data is mostly limited to skeletal muscle^5–7^ or blood plasma^8,9^ and randomized controlled trials are scarce and often characterized by small sample sizes. Given the broad implications of the immune system in protecting from disease, there is a clear rationale for comprehensive investigations of the impact of exercise on immune cells.

Both, the exercise-induced mobilization of specific immune cells into circulation as well as an increased cytokine release are well-described phenomena in exercise immunology. In detail, immune effector populations such as natural killer (NK) cells are highly sensitive to a mobilization by exercise^10,11^ and the humoral response is marked by the successive release of pro– and anti-inflammatory cytokines such as interleukin (IL)-6 and IL-10.^12^ While these physiological responses are well described, few investigations have evaluated the effect of acute exercise on molecular alterations from a cellular perspective. Applying bulk RNA sequencing to peripheral blood mononuclear cells (PBMCs), Contrepois et al. revealed a complex interplay of up– and downregulated transcripts that peaked two minutes after exercise and returned to baseline within 30-60 minutes. Enriched pathways were predominantly related to inflammatory signaling and immune activation, but also included other processes such as cell growth and mobility.^13^ While the number and kinetics of RNA transcripts suggest that many expected and some novel signaling pathways are initiated by acute exercise, the proteomic response of PBMCs remains unclear. Since the proteomic makeup of cells defines their phenotype and function, it is crucial to evaluate the impact of exercise on the cellular proteome of immune cells to draw informative conclusions about cell function. Therefore, we aimed to answer a simple but highly relevant question: does acute exercise alter the proteomic makeup of human immune cells, and if so, is this process dependent on exercise intensity?

To tackle this question, we tested two different endurance exercise paradigms that were chosen based on the World Health Organization 2020 guidelines on physical activity: high-intensity interval exercise (HIIE) and moderate-intensity continuous exercise (MICE). The guidelines recommend 150 – 300 minutes of moderate intensity or 75 – 150 minutes of vigorous intensity physical activity per week, as a result of accumulating evidence on the greater health benefits when exercising at higher intensities.^14^ Using state-of-the-art mass spectrometry-based proteomics in a standardized experimental setting, we demonstrate that the proteomic makeup of circulating immune cells is reshaped by acute exercise. Interestingly, proteomic alterations are more potently induced by HIIE compared to MICE. The observed changes indicate altered immune cell activation and effector functions in the recovery phase following HIIE, which underlines the immunomodulatory impact of exercise on a proteomic level. Of note, flow cytometry-based immune cell phenotyping suggests that these adaptions occur independent of immune cell mobilization, as indicated by identical mobilization patterns between HIIE and MICE. Using prediction models, we additionally identify an immunoproteomic signature associated with cardiorespiratory fitness. We provide a highly reliable and comprehensive data resource on the impact of acute exercise on immune cell proteomes, including repeated baseline measures and exercise responses in humans. Our data indicates that exercising at higher intensities is necessary to induce proteomic changes associated with immune function, providing biological support for exercise intensity as a crucial variable in the WHO 2020 physical activity guidelines.

## RESULTS

### Study design and participant characteristics

To investigate the impact of exercise intensity on the proteome of immune cells in a standardized experimental setting, we designed a randomized crossover study comparing time– and workload-matched HIIE with MICE. The study was prospectively registered at the German Clinical Trials Register (DRKS00017686) and met the National Institutes of Health’s definition of a clinical trial. In total, data from 23 overnight-fasted recreationally active runners (12 female, 11 male) was collected during three visits. Participants exhibited a mean (±SD) age of 30 ± 4 years, a body mass index (BMI) of 22.2 ± 2.38 kg m^-2^, and a cardiorespiratory fitness (measured as peak oxygen uptake, VO2peak) of 56.64 ± 6.43 mL min^-1^ kg^-1^.

To assess VO2peak and determine individual exercise intensities for the subsequent HIIE and MICE sessions, participants performed a cardiopulmonary exercise test (CPET) on a treadmill until volitional exhaustion on their first visit. Respiratory exchange ratio (RER) at termination of the CPET was 1.07 ± 0.05. Participants were subsequently randomized into one of two intervention sequences, which did not differ in terms of participant characteristics, indicating that our randomization was unbiased (Table S1). According to their randomization sequence, participants then performed HIIE and MICE on separate days. Both exercise interventions were performed on a treadmill and matched for exercise duration and workload^15,16^ to isolate exercise intensity as an independent factor. Although often disregarded in exercise research, this time– and workload-matched design is crucial to draw unbiased conclusions on the impact of exercise intensity. PBMCs were isolated at baseline, immediately after, and 1h after exercise (Figure 1A).

**Figure 1.**
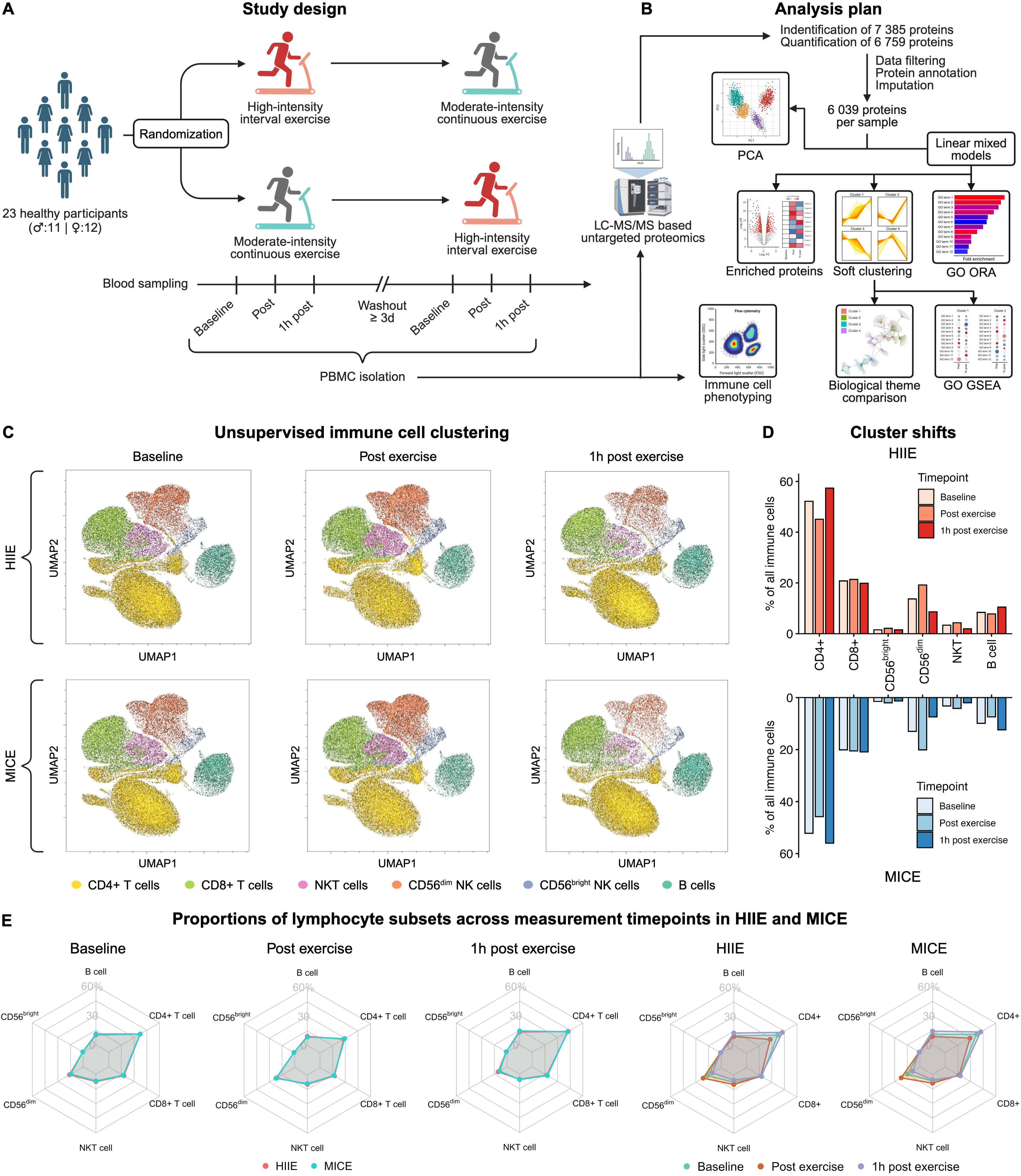
Study design, analysis plan, and exercise-induced immune cell mobilization. (A) Overview of the study design including time– and workload-matched high-intensity interval exercise (HIIE) and moderate-intensity continuous exercise (MICE). (B) Overview of the bioanalytical and bioinformatic methods used to analyze peripheral blood mononuclear cells (PBMCs). (C) Uniform Manifold Approximation and Projection (UMAP) of immune cell clusters identified by unsupervised clustering using self-organizing maps (SOM). Immune cell clusters are displayed color-coded and separated by exercise condition, and measurement timepoint. Each UMAP corresponds to 3,000 vital lymphocytes from 22 samples, resulting in a total of 66,000 events. For the UMAP representing 1h post MICE only 21 samples were available, resulting in 63,000 events. (D) Comparison of exercise-induced shifts in the identified clusters. (E) Proportions of lymphocyte subsets in response to HIIE and MICE. See also Figure S1 and Tables S2, S3, and S4.

After termination of the study, the collected samples were analyzed via two different bioanalytic pipelines: (1) state-of-the-art liquid-chromatography mass-spectrometry-based proteomics using data-independent acquisition for identification and quantification of intracellular alterations in protein abundance, and (2) flow-cytometry based immune cell phenotyping to assess exercise-induced changes in circulating immune cell numbers. Our proteomics analysis yielded an excellent coverage with a total of 7,385 identified and 6,759 quantified proteins in the whole dataset, which is comparable to other studies in the field of immunology. Of note, these studies often use murine immune cells^17^ or resting samples from healthy donors,^18^ making our study the first to apply large-scale proteomic analyses in a randomized interventional setting with repeated baselines. Data filtering, protein annotation, and imputation of missing values resulted in 6,039 proteins across 23 participants in 2 exercise conditions with 3 measurement timepoints, respectively (for details see methods). This makes our dataset the largest immune cell proteomics data source available in exercise context to date (Figure 1B).

To account for a potential bias in our proteomics analysis arising from an exercise-induced shift within the PBMC compartment, we additionally performed immune cell phenotyping by flow cytometry (Figure S1A). So far this has been disregarded in studies applying omics approaches on immune cells in exercise context.^13^ Flow cytometry yielded a total of 3,537,855 vital lymphocytes across all samples with a mean of 27,007 ± 7,956 analyzed cells per sample (Table S2).

### Immune cell mobilization and redistribution is independent of exercise intensity

The mobilization and redistribution of immune cells in response to acute exercise is one of the core phenomena of exercise immunology and it is nowadays agreed upon that the recovery phase following exercise is characterized by a transmigration of lymphocytes from the bloodstream into peripheral tissues, with crucial implications in many disease settings, including anticancer immunity,^19,20^ and immunological defense.^21,22^ A remaining topic of debate, however, is whether exercise intensity influences the magnitude of immune cell mobilization since previous studies on this topic were matched for exercise duration, but not workload.^23,24^ Thus, before dissecting proteomic alterations of PBMCs in response to exercise, we aimed to clarify whether exercise intensity has an impact on immune cell mobilization and redistribution, since this would lead to a different composition of our PBMC samples in response to HIIE and MICE.

To answer this question, we performed spectral flow cytometry on all samples and applied unsupervised immune cell clustering using self-organizing maps (SOM) with 3,000 vital lymphocytes per sample. This was done with downsampled data to maintain the proportions of the underlying immune cell populations. Using Uniform Manifold Approximation and Projection (UMAP), we identified 6 separate clusters, which were mapped to the corresponding immune cell populations by comparison to the fluorescence intensities of antibody-tagged target proteins. Visual inspection of the generated UMAPs suggested alterations in the density of immune cell clusters by exercise without considerable differences between HIIE and MICE (Figure 1C). Quantification of exercise-induced cluster shifts resulted in a similar distribution pattern of immune cell clusters between HIIE and MICE with a mean delta of 0.004 ± 0.9 % (Figure 1D; Table S2). This suggest that HIIE and MICE induce similar mobilization patterns, independent of exercise intensity.

To confirm these results, we additionally determined absolute numbers of circulating lymphocytes and quantified exercise-induced alterations of all immune cell populations. Linear mixed models yielded significant alterations over time for all lymphocyte subsets except regulatory T cells. Of note, there were no significant time × condition interaction effects, suggesting that immune cell mobilization and redistribution was identical between HIIE and MICE (Figure S1B; Table S3). We also determined the proportional contribution of each immune cell population to the PBMC compartment to assess the impact of exercise on potential shifts in PBMC composition. As expected, PBMC proportions were altered by exercise, but did not differ between HIIE and MICE (Figure 1E; Table S4).

Collectively, using an unsupervised clustering approach and a conventional statistical approach, we demonstrate that HIIE and MICE cause identical kinetics with respect to immune cell mobilization and redistribution when matched for exercise duration and workload. This proves that intensity *per se* does not impact these processes. The homogeneous shift in PBMC composition between HIIE and MICE confirmed these findings and served as crucial prerequisite for the subsequent analysis of alterations in the PBMC proteome.

### Measures of variability indicate high reliability of the generated proteomics dataset

Before conducting statistical analyses of exercise-induced alterations to the immune cell proteome, we evaluated the analytical quality of our generated dataset. Inter-individual variability of all quantified proteins resulted in a median coefficient of variation (CV) of 2.56 (IQR = 1.81), 2.82 (IQR = 2.03), and 3.15 (IQR = 2.20) for baseline, post exercise and 1h post exercise in HIIE, and a median CV of 3.14 (IQR = 2.33), 2.91 (IQR = 2.41), and 2.95 (IQR = 2.34) for the same measurement timepoints in MICE (Figure 2A). Of note, these CVs are considerably lower than in other proteomics studies in exercise context,^13,25^ which underlines the homogeneity of our study population and the analytic quality of our proteomics pipeline. Given that each participant performed two different exercise interventions on separate days, the applied crossover design additionally enabled us to obtain repeated baseline measurements and calculate intra-individual variability (i.e., protein variability within each participant). The mean difference between the two baselines amounted to 0.13 ± 0.75 % for females and 0.06 ± 0.59 % for males (Figure 2B). To assess variability on a per-protein level, we combined multiple measures of variability (mean CV at baseline, mean CV in response to exercise, and mean difference between the two baselines) into an integrated variability score. 99.34 % of all quantified proteins revealed a proteomic variability of < 10 % and 83.34 % of all proteins achieved a score < 5 % (Figure 2C, S2A, and S2B). In summary, the low variability emphasizes the high quality of our study setup and proteomics workflow and proves that the proteomics dataset we generated is highly reliable.

**Figure 2.**
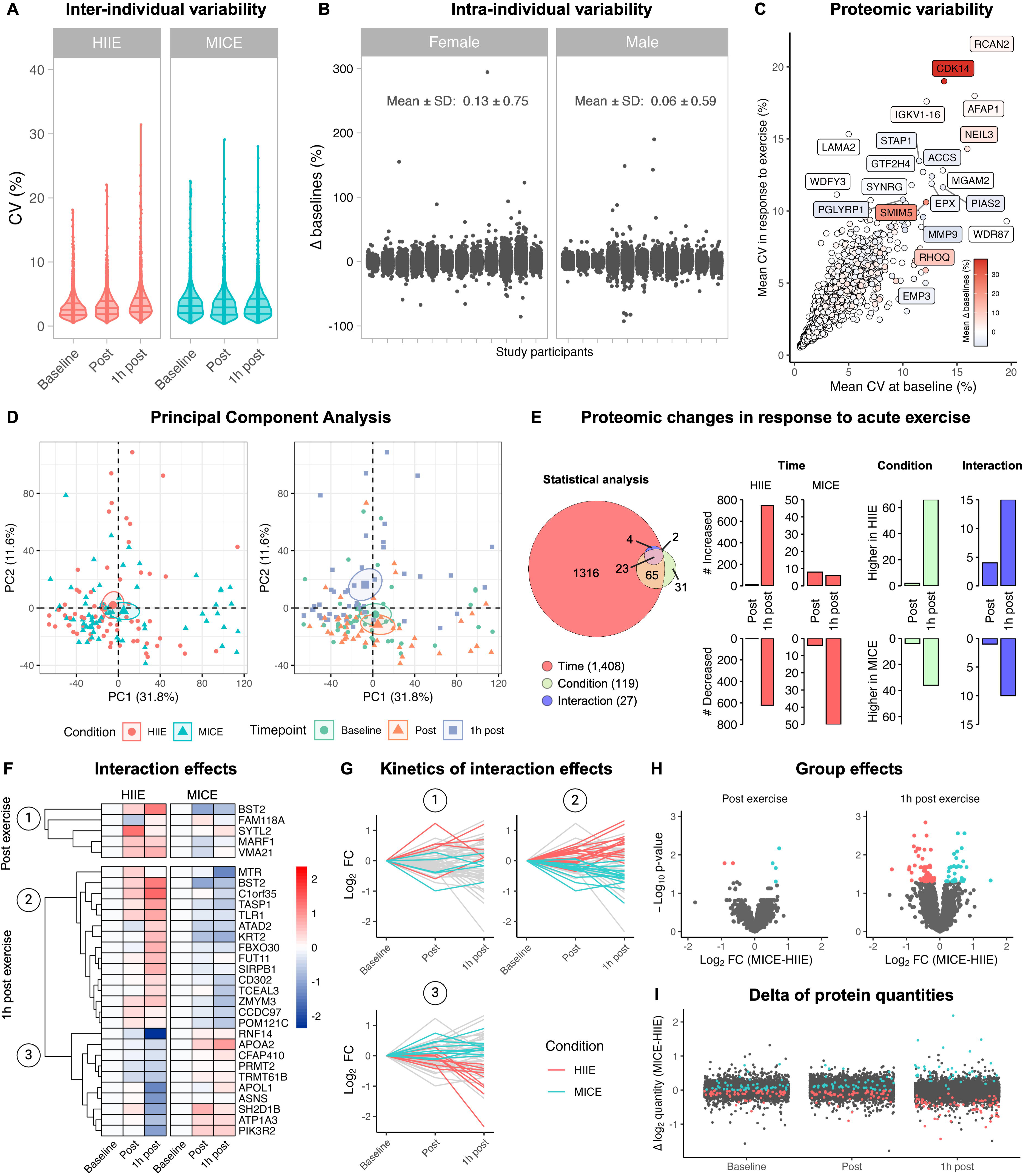
Measures of variability and proteomic changes in response to exercise. (A) Inter-individual variability of all quantified proteins expressed as coefficients of variation (CVs) separated by exercise condition and timepoint. (B) Intra-individual variability of all quantified proteins expressed as delta between the two baselines separated by sex and study participant. (C) Proteomic variability of all quantified proteins. (D) Principal component analysis of all samples using exercise condition and measurement timepoint as metadata. Small symbols indicate individual samples. Big symbols and circles indicate mean and 95 % confidence interval of the corresponding data subset. (E) Quantification of proteomic changes in response to exercise. Linear mixed models containing exercise condition, measurement timepoint, and the interaction between both as fixed factors, were applied (n = 23). (F) Interaction effects between time and exercise condition for HIIE and MICE. Dendrograms depict clusters identified by full-linkage hierarchical clustering. (G) Kinetics of proteins displaying interaction effects separated by the identified clusters (1-3). (H) Group differences between HIIE and MICE. Significant proteins are colored by exercise condition. (I) Delta of protein quantities between HIIE and MICE. Significant proteins are colored by exercise condition. See also Figure S2 and Table S5.

### Acute exercise alters the immune cell proteome

To obtain first insights into the proteomic alterations induced by exercise, we performed principal component analysis (PCA) with all collected samples. Visual inspection of the PCAs suggested that the variation within our samples was mainly accounted for by measurement timepoints (i.e., baseline, post exercise, 1h post exercise), but not exercise condition *per se*, as indicated by a separated 95 % confidence interval (CI) 1h after exercise (Figure 2D). PCA also suggested that sex and intervention day had little impact on the variation of our data (Figure S2C). Building upon these results, we additionally performed PCAs on subsets of our data and observed a separated 95 % CI for the same measurement timepoint (1h post) when performing PCA only with HIIE samples. Of note, this effect was not observed when using MICE samples, suggesting that the recovery phase after HIIE accounts for more variation in our samples compared to MICE (Figure S2D). A similar tendency for the 1h post measurement timepoint was observed when performing PCAs separated by timepoint (Figure S2E).

Using linear mixed models, we next compared the impact of HIIE and MICE on proteomic alterations in PBMCs. We identified 1,408 time effects, 119 group effects, and 27 time × group interaction effects (Figure 2E). Including sex as a fixed effect in our linear mixed models did not result in any statistically significant results. Dissection of the obtained results uncovered that there were more time effects in HIIE compared the MICE, and more group effects 1h after exercise compared to immediately after exercise. Time × group interaction effects also showed more differences between HIIE and MICE 1h after exercise compared to immediately after (Figure 2E). This is in line with our results obtained by PCA and proves that the immune cell proteome is more severely altered by HIIE compared to MICE. In detail, HIIE was marked by 1,377 significantly altered proteins, while MICE only caused significant alterations in 64 proteins. The higher number of time effects 1h after exercise compared to immediately after exercise may result from the fact that this measurement timepoint falls within a timeframe that is sufficient for transcription and translation to take place.^26^ Apart of *de novo* synthesis of proteins, there are of course also other mechanisms at play such as endocytosis and exocytosis, protein degradation, or extracellular vesicle-mediated uptake and release.

#### Proteomic alterations differ between HIIE and MICE

The fact that our study was designed to identify intensity-dependent differences in proteomic alterations over time, emphasizes the statistical power of the observed time × group interaction effects. Immediately after exercise, we observed 5 proteins with distinct kinetics in HIIE compared to MICE (Figure 2F). Among these proteins, synaptotagmin-like protein 2 (SYTL2), a crucial contributor to cytotoxic granule exocytosis in lymphocytes, displayed a strong increase in response to HIIE, while it remained unaltered in MICE. Similarly, bone marrow stromal antigen 2 (BST2) – known for its role in blocking virus release from infected cells – increased in response to HIIE but decreased in MICE (Figure 2F and 2G). This gives first insights into the immunomodulatory potential of HIIE and suggests immunological adaptions dependent on exercise intensity already immediately after exercise.

Building upon these results, we observed 25 interaction effects in the recovery phase after exercise. Hierarchical clustering yielded two major clusters of proteins marked by opposed kinetics in HIIE compared to MICE (Figure 2F and 2G). For instance, BST2, toll-like receptor 1 (TLR1) and cluster of differentiation 302 (CD302) were increased 1h after HIIE but decreased in MICE. TLR1 is the most abundantly expressed TLR on NK cells^27^ and serves as membrane-bound pattern recognition receptor for microbial lipopeptides. Upon stimulation, TLRs trigger cytokine production and NK cell cytotoxicity to combat viral and bacterial infections.^28,29^ Several studies have demonstrated that this mechanism is dependent on TLR1,^30,31^ suggesting that exercise-induced increases in TLR1 might reinforce NK cell-mediated immunity against invading pathogens. Similarly, C-type lectin receptors play a crucial role in innate immunity and inflammatory responses.^32,33^ On macrophages colocalization of F-actin with the C-type lectin receptor CD302 was shown to contribute to cell adhesion and migration.^34^ Thus, exercise-induced increases in CD302 might render circulating immune cells more susceptible to tethering and rolling on the endothelium followed by transendothelial migration and infiltration into peripheral tissues. Of note, BST2 was the only protein that continued to increase from post exercise to 1h post exercise in HIIE, suggesting sustained intensity-dependent adaptions in antiviral defense. As a core host restriction factor, increased BST2 levels in response to acute bouts of high-intensity exercise might result in chronic adaptions of antiviral immunity, thus providing a potential link to the reduced infection risk, disease severity and mortality observed in individuals performing high levels of physical activity.^35–37^

In contrast, proteins such as asparagine synthetase (ASNS) or SH2 domain-containing protein 1B (SH2D1B) were marked by a decrease in the recovery period following HIIE, while they remained unaltered or increased in MICE (Figure 2F and 2G). ASNS plays a crucial role in asparagine biosynthesis and was previously shown to regulate CD8+ T cell activation, differentiation, and effector function.^38,39^ Since these processes are regulated by a fine-tuned equilibrium between intracellular biosynthesis versus cellular uptake of asparagine, our data suggests less reliance on de novo synthesis in circulating immune cells. These results are underlined by reduced plasma levels of asparagine in response to both, endurance and resistance exercise,^8^ making cellular uptake of asparagine by circulating immune cells a tempting hypothesis. Similarly, SH2D1B, a cytoplasmic adapter regulating NK cell effector functions, was differentially expressed between HIIE and MICE 1h after exercise.

Finally, our statistical analysis also yielded several group differences between HIIE and MICE. As for time and interaction effects, we observed more pronounced group differences 1h after exercise compared to immediately after exercise (Figure 2H). Calculation of the differences in protein quantity between HIIE and MICE confirmed this response as demonstrated by increasing deltas for these proteins from baseline to 1h post exercise (Figure 2I). Detailed statistical results for all proteins can be explored in Table S5. In summary, our results obtained by PCA and linear mixed models prove that the recovery phase following HIIE is marked by more profound alterations to the immune cell proteome compared to MICE. Additionally, we provide evidence that several proteins related to immune effector functions are differentially expressed over time between HIIE and MICE. Against the backdrop of our flow cytometry results, these effects occur despite identical immune cell mobilization between the two exercise conditions.

### Exercise reshapes the immune cell proteome towards effector function

Considering the high number of time effects compared to group and interaction effects, we next aimed to dissect the proteomic alterations over time. To add a functional dimension to these time effects, we made use of the Gene Ontology (GO) Resource.^40,41^ GO over-representation analysis yielded 27 enriched GO terms in HIIE and 9 enriched GO terms in MICE. Interestingly, enriched GO terms were centered around immune effector functions in both HIIE and MICE with biological processes like “disruption of cell in another organism” or “killing of cells of another organisms” yielding high fold enrichments and – log_10_-transformed p-values. A further biological process related to immune effector function that was shared between HIIE and MICE was “cell killing” (Figure 3A).

**Figure 3.**
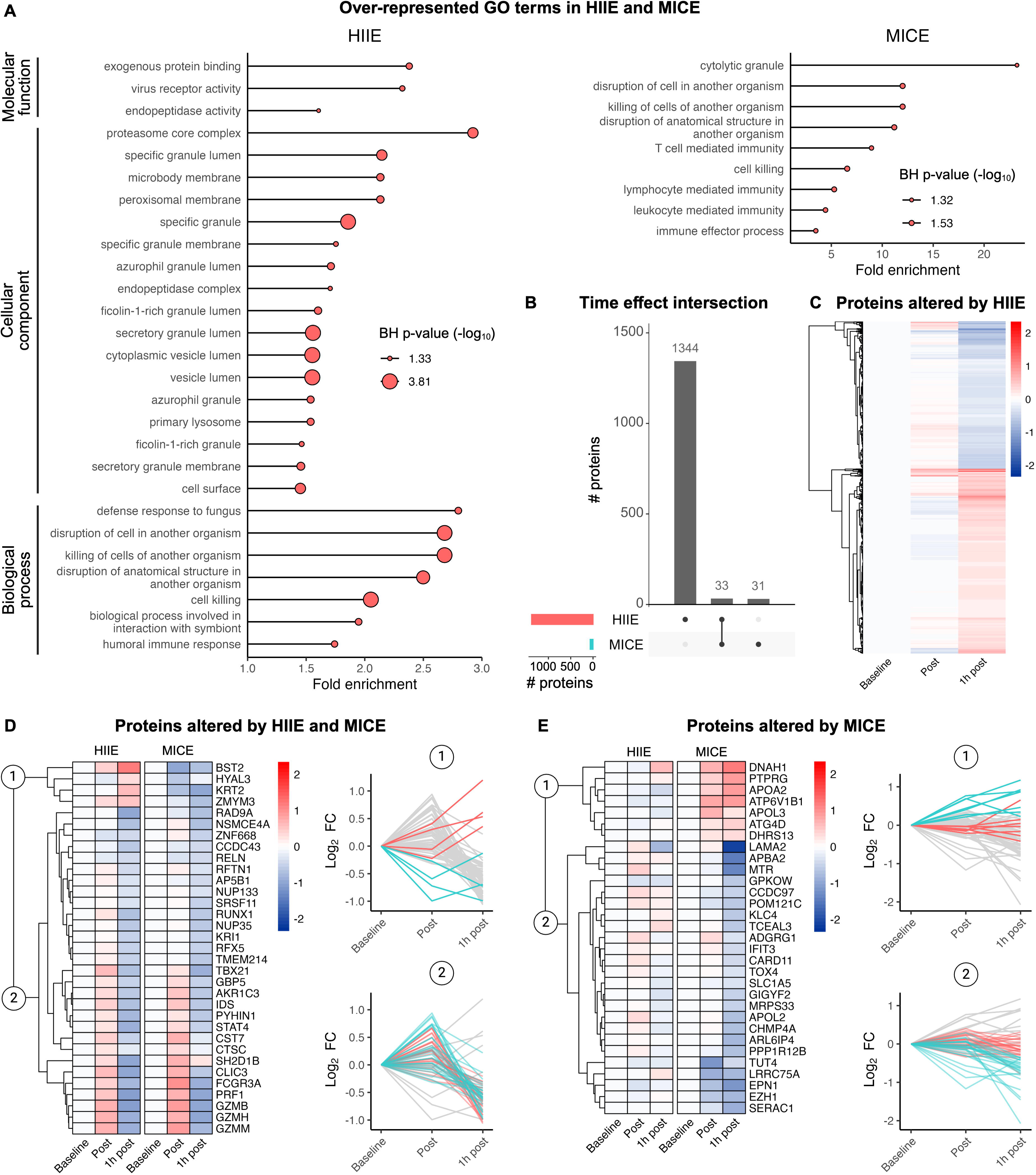
Exercise reshapes the immune cell proteome towards effector function and causes distinct alterations in protein abundance in response to HIIE and MICE. (A) Gene ontology (GO) over-representation analysis comparing significantly altered proteins in HIIE (n = 1,377) and MICE (n = 64) with all proteins quantified in this study (n = 6,039). (B) Overview of proteins altered by HIIE, MICE, or both exercise conditions. (C) Overview of proteins uniquely altered by HIIE. (D) Overview of proteins altered by both exercise conditions. The first two clusters identified by hierarchical clustering separate proteins displaying different kinetics in HIIE and MICE (1) from proteins displaying similar kinetics (2). (E) Overview of proteins uniquely altered by MICE. The first two clusters identified by hierarchical clustering separate proteins increasing in response to MICE (1) from proteins decreasing in response to MICE (2). See also Table S5 and S6.

For proteins altered by MICE, GO over-representation analysis additionally yielded several biological processes related to lymphocyte effector function, such as “lymphocyte mediated immunity” or “T cell mediated immunity”. Given that the over-representation analysis was conducted with much more proteins for HIIE, we additionally identified several cellular components and molecular functions in this analysis. Semantic evaluation of the identified GO terms underlined their association with immune effector function. For instance, “exogeneous protein binding”, “virus receptor activity”, and “endopeptidase activity” are known molecular functions in the context of immunological defense against viruses.^42,43^ Similarly, “proteasome core complex” and “peroxisomal membrane” depict cellular components associated with such molecular function and biological processes (Figure 3A). Interestingly, some cellular components also suggested effector functions such as vesicle-mediated lysis of neighboring cells, e.g., in the context of tumor killing.^44,45^ Examples for such cellular components are “cytoplasmic vesicle lumen” or “secretory granule lumen” (Figure 3A).

Collectively, our GO over-representation analysis points towards enhanced regulation of immune effector functions in response to both HIIE and MICE. Detailed results of the over-representation analysis can be explored in Table S6.

### Time-resolved protein changes differ between HIIE and MICE

To comprehensively assess the regulation of immune effector functions in response to exercise, we next evaluated temporal alterations in protein abundance in response to HIIE and MICE. Within all proteins altered by exercise (n = 1,408), we found 1,344 proteins that were uniquely altered by HIIE, 31 proteins that were uniquely altered by MICE, and 33 proteins that were altered by both exercise conditions (Figure 3B). Analysis of proteins altered by HIIE suggested two major protein clusters that were characterized by increased or decreased protein abundance 1h after HIIE compared to baseline (Figure 3C). Considering the large number of proteins altered by HIIE compared to MICE, we took different approaches in analyzing the time effects of each exercise condition.

For proteins altered by both exercise conditions, and proteins uniquely altered by MICE we performed hierarchical clustering to identify proteins displaying similar kinetics. Interestingly, when analyzing proteins that were altered by both exercise conditions, hierarchical clustering yielded 2 major protein clusters: one cluster containing proteins with similar kinetics between HIIE and MICE and on cluster containing proteins with different kinetics (Figure 3D). In absolute terms, most of the proteins responded similarly with only 4 proteins showing higher values in HIIE, including the antiviral protein BST2, which we previously identified in our analysis of time × group interaction effects (Figure 2F). Additionally, many of the proteins that were shared between HIIE and MICE were associated with immune effector functions, suggesting a shared regulation of several immunological processes by exercise. Examples of such proteins include granzymes (e.g., GZMB, GZMH, GZMM), perforin-1 (PRF1), T-box transcription factor 21 (TBX21), or guanylate-binding proteins 5 (GBP5; Figure 3D).

Similarly, hierarchical clustering of proteins uniquely altered by MICE identified 2 major clusters that separated proteins that decreased in response to MICE from proteins that increased, while showing no alterations in HIIE, respectively (Figure 3E). In line with the observed time × group interaction effects (Figure 2F) most proteins displayed lower abundance in response to MICE. Protein candidates underlining this are the translational repressor GRB10-interacting GYF protein 2 (GIGYF2), the signal transducer caspase recruitment domain-containing protein 11 (CARD11) and the antiviral protein interferon-induced protein with tetratricopeptide repeats 3 (IFIT3; Figure 3E). Of note, although the lower number of time effects and the decreased abundance of many proteins might suggest reduced immune effector functions in response to MICE, it is crucial to emphasize that several proteins with immunological functions, especially those jointly regulated between HIIE and MICE, revealed increased abundance in response to MICE as well. Thus, while our data suggests that the immunoproteomic impact of MICE seems to be less pronounced than that of HIIE, there is no conclusive evidence suggesting reduced immune effector function *per se* in response to MICE.

### Biological theme comparison suggests shared and unique GO terms across protein clusters in HIIE

While hierarchical clustering proved powerful to identify shared protein kinetics in MICE (Figures 3D and 3E), it does not offer evaluation of the relative membership of proteins to different identified clusters. Thus, to map proteins altered by HIIE to distinct clusters by means of their relative membership, we performed fuzzy c-means clustering on all proteins altered by HIIE (n = 1,377). We identified 4 different protein clusters (Figure 4A; Table S7), which confirmed what hierarchical clustering had previously suggested, i.e., two major clusters marked by increased or decreased protein abundance 1h after HIIE (Figure 3C). Building upon this, the 4 identified clusters demonstrated different protein kinetics with mean membership values of 58 %, 67 %, 66 %, and 60 % for the underlying proteins of clusters 1 to 4, respectively (Figure 4A). Inspection of the relative membership across all clusters suggested that most proteins were clearly mapped to a specific cluster with only few proteins scoring high or intermediate in more than one cluster (Figure 4B). This suggests that our approach was successful in identifying different clusters within proteins altered by HIIE. The identified clusters were taken forward in our analytic workflow to analyze the functional consequences of proteomic alterations in response to HIIE.

**Figure 4.**
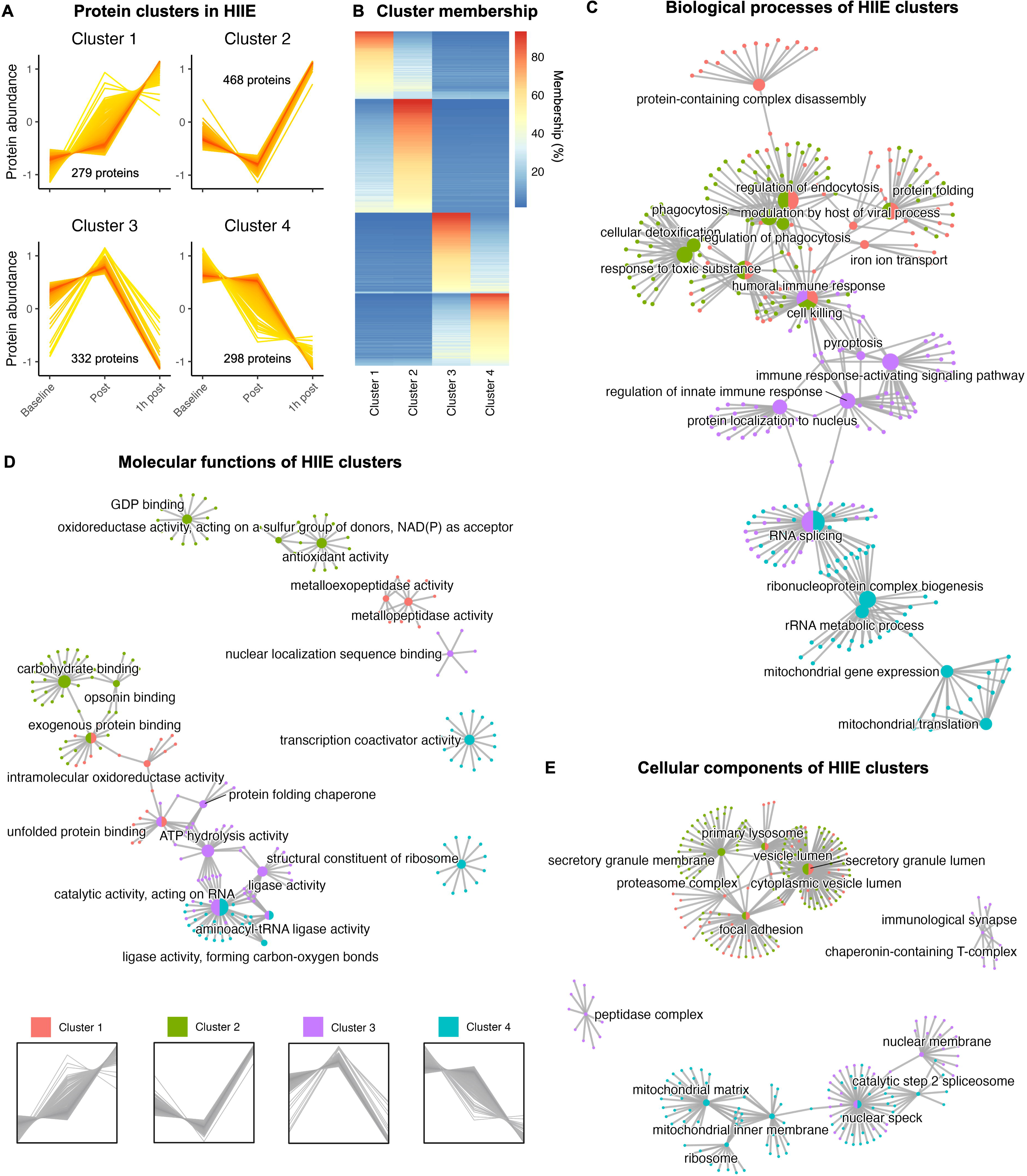
Biological theme comparison suggests shared and unique GO terms across protein clusters in HIIE. (A) Protein clusters identified by fuzzy c-means clustering in response to HIIE. (B) Relative cluster membership of proteins altered by HIIE. (C) Comparison of biological processes in clusters 1 – 4. (D) Comparison of molecular functions in clusters 1 – 4. (E) Comparison of cellular components in clusters 1 – 4. See also Table S7 and S8.

We next linked the observed proteomic alterations to cellular processes to gain insights into the functional consequences for immune cells. To accomplish this, we leveraged biological theme comparisons.^46^ We identified 576 biological processes, 132 molecular functions, and 187 cellular components associated with the proteins altered by HIIE and observed various shared and unique GO terms across protein clusters 1 – 4 (Table S8). To unravel the biological implications of these results, we generated gene-concept networks of the 5 most significant GO terms in each ontology (Figure 4C – E).

Regarding biological processes, “cell killing” was shared between clusters 1 – 3, while “regulation of endocytosis” and “humoral immune response” were only shared between clusters 1 and 2. In contrast, clusters 3 and 4 shared many proteins annotated to “RNA splicing” and all clusters yielded GO terms that were uniquely identified in one cluster, respectively (Figure 4C). Interestingly, semantic analysis of shared and unique molecular functions confirmed these results: cluster 1 and 2 shared “exogeneous protein binding” as a molecular function, which complements the biological process “regulation of endocytosis”. Similarly, clusters 3 and 4 shared “catalytic activity, acting on RNA”, which complements the biological process “RNA splicing” (Figure 4D). As expected, these similarities were also mirrored on the level of cellular components. While clusters 1 and 2 shared “focal adhesion” and “secretory granule lumen”, clusters 3 and 4 shared “nuclear speck”, a subnuclear structure known to contain large amounts of splicing factors (Figure 4E).

Although the semantic interconnection of these biological processes, molecular functions, and cellular components is strongly driven by the annotation of similar underlying proteins to each of the GO terms, respectively, the biological theme comparison of clusters 1 – 4 emphasized a crucial aspect of our analysis: despite considerable differences in the temporal kinetics of the underlying proteins, there seem to be shared processes that are regulated irrespective of the underlying protein kinetic. In contrast, some processes are only regulated by proteins displaying a specific kinetic, i.e., proteins belonging to a specific cluster. We thus next aimed to comprehensively dissect the regulated GO terms, to identify shared and unique biological processes, molecular functions, and cellular components across protein clusters 1 – 4.

### Identification of core immune effector functions regulated by HIIE

Building upon our results obtained by over-representation analysis and biological theme comparison, we next aimed to gain insights into the direction and magnitude of regulated biological processes, molecular function, and cellular components. We thus performed gene set enrichment analyses (GSEA),^47^ which in addition to the underlying proteins also takes the fold changes from baseline – in our case induced by HIIE – into account. This depicts a valuable extension to our previous analyses (Figure 3A and 4C – E) since it adds a quantitative measure of enrichment (normalized enrichment score; NES) to our results.

GSEA yielded a total of 169 enriched GO terms, which were in part shared between different clusters, and in part unique to specific clusters (Figure 5A and Table S9). Interestingly, cluster 4 did not yield any enriched GO terms and re-evaluation of the underlying statistics demonstrated that the individual terms were all below the significance threshold. To validate these findings, we ran functional enrichment analyses using the STRING resource^48^ and obtained the same results with no enriched GO terms for cluster 4. Before analyzing the functional implications of cluster-specific GO terms, we focused our attention on GO terms that were shared across different protein clusters. In total, 11 biological processes were shared across protein clusters 1 – 3, suggesting a shared regulation by HIIE, independent of the underlying protein kinetics. Semantic evaluation demonstrated a close connection to immune function, as exemplified by GO terms like “cell killing”, “leukocyte activation”, or “defense response” (Figure 5A). Analysis of the underlying proteins that were annotated to these GO terms resulted in a core proteome consisting of 369 proteins, most of which changed in abundance in the recovery phase following HIIE (Figure 5B). This suggests that the biological processes regulated by HIIE are driven by proteomic alterations in the recovery phase following HIIE. Of note, although these biological processes revealed increased regulation in some clusters, while demonstrating decreased regulation in others – a finding that is easily explained by different underlying proteins – the conclusion remains unchanged: these core immune effector functions are regulated by HIIE independent of the underlying protein kinetics. Similarly, we observed 27 GO terms that were shared between clusters 1 and 2, and 15 GO terms that were shared between clusters 2 and 3 (Figure 5A). Here, analysis of the underlying proteins yielded a core proteome consisting of 528 and 188 proteins, most of which also strongly increased in the recovery phase following HIIE (Figure S3A-C). Core biological processes regulated by these proteins included “cytokine production” and “secretory vesicle” for clusters 1 and 2 (Figure S3B), and “leukocyte migration”, “cell-cell adhesion” and “regulation of lymphocyte activation” for clusters 2 and 3 (Figure S3C). A detailed overview of the direction and magnitude of all regulated biological processes, molecular functions, and cellular components shared between protein clusters 1 – 3 is given in Figure S4.

**Figure 5.**
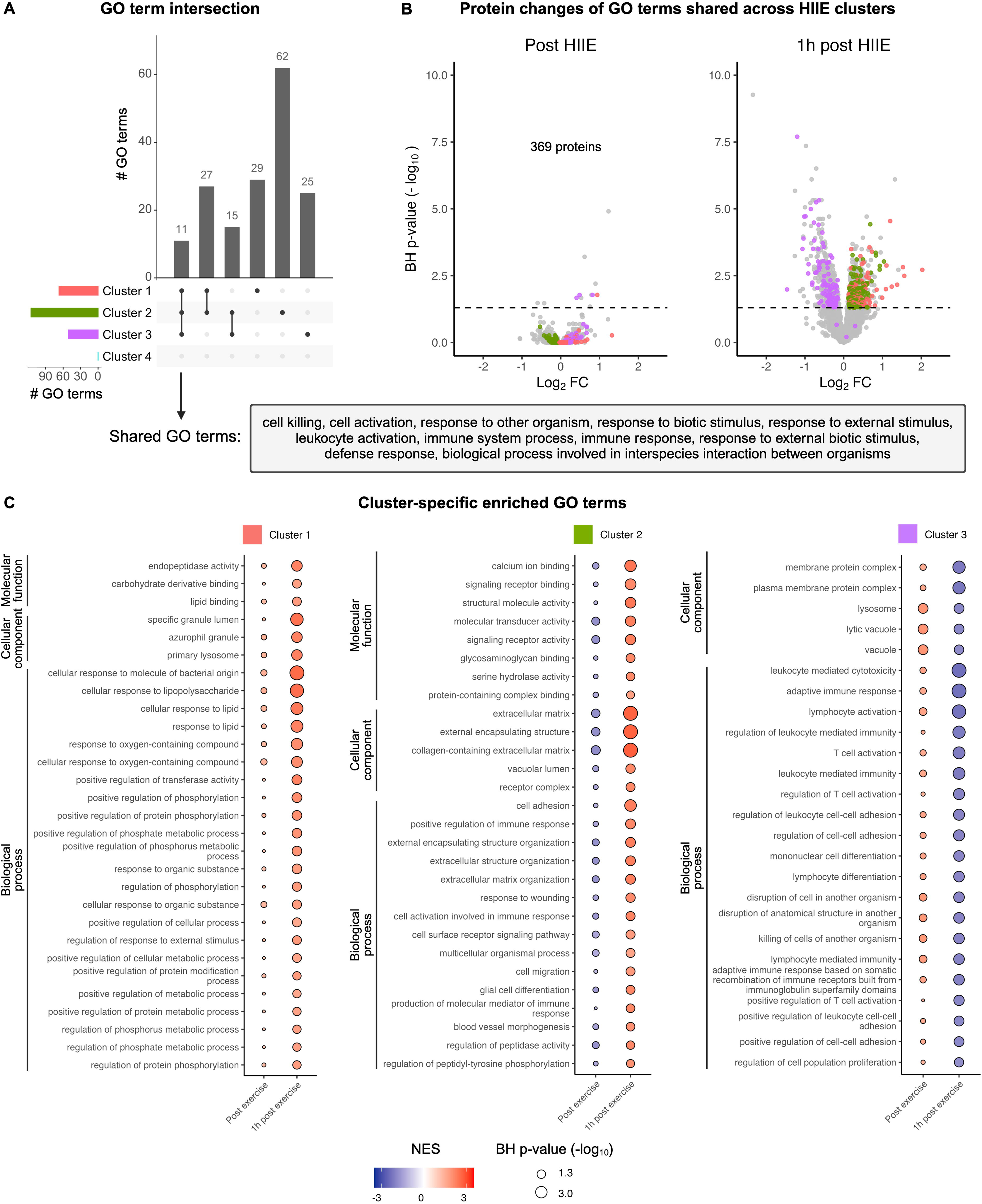
Identification of core and cluster-specific immune effector functions in the recovery phase following HIIE. (A) Shared and unique GO terms across protein clusters 1 – 3. (B) Identification and temporal regulation of core biological processes and the underlying proteins regulated by clusters 1 – 3. (C) Cluster-specific GO terms regulated by HIIE. See also Figure S3, S4, and S5 and Table S9.

### Identification of cluster-specific immune effector functions

Having identified core biological processes regulated by HIIE, we next focused our attention on GO terms that were uniquely enriched in a specific protein cluster. This approach enabled us to gain a better understanding of the magnitude and direction of biological processes that were driven by proteins displaying a certain kinetic. We identified 29 GO terms uniquely enriched in cluster 1, 62 GO terms uniquely enriched in cluster 2, and 25 GO terms uniquely enriched in cluster 3 (Figure 5A).

Among the GO terms enriched in cluster 1 we found enriched regulation of “endopeptidase activity” and “cellular response to organic substance”, suggesting higher regulation of mechanisms associated with immunological defense in response to HIIE (Figure 5C). Similarly, cluster 2 demonstrated enriched regulation of “glycosaminoglycan binding”, “cell migration”, and “production of molecular mediator of immune response”, which underlines the immunological implications of proteins associated with cluster 1 and 2. As previously demonstrated, the enriched regulation of cellular processes was strongly reserved to the recovery phase following HIIE with only few enriched GO terms immediately after exercise. Cluster 2 also revealed enriched regulation of several components of immune cell receptor signaling and downstream signal transduction, including “cell activation involved in immune response”, “cell surface receptor signaling pathway”, and “molecular transducer activity”. Of note, cluster 1 and 2 also contained several proteins that were explicitly annotated to have a positive impact on biological processes, thereby partially overcoming the lack of directionality that GO analyses are usually characterized by. For instance, we observed a positive regulation of protein phosphorylation, transferase activity, and metabolic processes in cluster 1, and a positive regulation of immune response in cluster 2 (Figure 5C).

In contrast, cluster 3 was characterized by a decreased regulation of several cellular components and biological processes 1h after exercise, which is easily explained by the underlying protein kinetic. While clusters 1 and 2 were marked by an increased protein abundance 1h after HIIE, cluster 3 demonstrated a decreased protein abundance for the same measurement timepoint (Figure 4A). To resolve this discrepancy arising from contrary protein kinetics, we re-ran GSEA with all proteins altered by HIIE and mined the results for highly regulated biological processes, molecular functions and cellular components. Interestingly, this global analysis yielded similar results to our cluster-specific analysis (Figure S5A-C). Detailed results of the global GSEA can be explored in Table S9. In summary, our GSEA suggested a profound regulation of immune effector processes in the recovery phase following HIIE. These processes were in part shared and in part unique for specific protein clusters. A remaining limitation, however, is the limited interpretation with respect to directionality. To overcome this, biological background information on the proteins annotated to specific GO terms must be integrated into the observed protein kinetics.

### Identification of an immunoproteomic signature associated with cardiorespiratory fitness

Having characterized the proteomic response of circulating immune cells to acute exercise, we ultimately leveraged our generated dataset to enable deeper insights into long-term adaption processes to exercise training. Taking an unbiased approach to this, we started by pooling the baseline data of all our analyses, including sex, anthropometric data, flow cytometry-based immunophenotyping, and mass spectrometry-based proteomics. We then used this comprehensive dataset to investigate potential pairwise associations with VO2peak, a gold standard marker of cardiorespiratory fitness that is highly responsive to exercise training. In a first step we calculated Spearman’s rank correlation coefficients (r_S_) and selected features that displayed moderate to high correlation (r_S_ > 0.4 or < –0.4) with VO2peak. This resulted in a reduction of our dataset from 6,063 to 260 features.

To establish an elaborate connection between these features and cardiorespiratory fitness, we next performed prediction analyses to gain a better understanding of the magnitude and direction of their impact on VO2peak. Ridge regression yielded an R-squared (R^2^) of 0.61 and a mean squared error (MSE) of 14.1 and visual inspection of the ranked coefficients revealed a homogeneous distribution of features with positive and negative impact on VO2peak prediction, respectively (Figure 6A).

**Figure 6.**
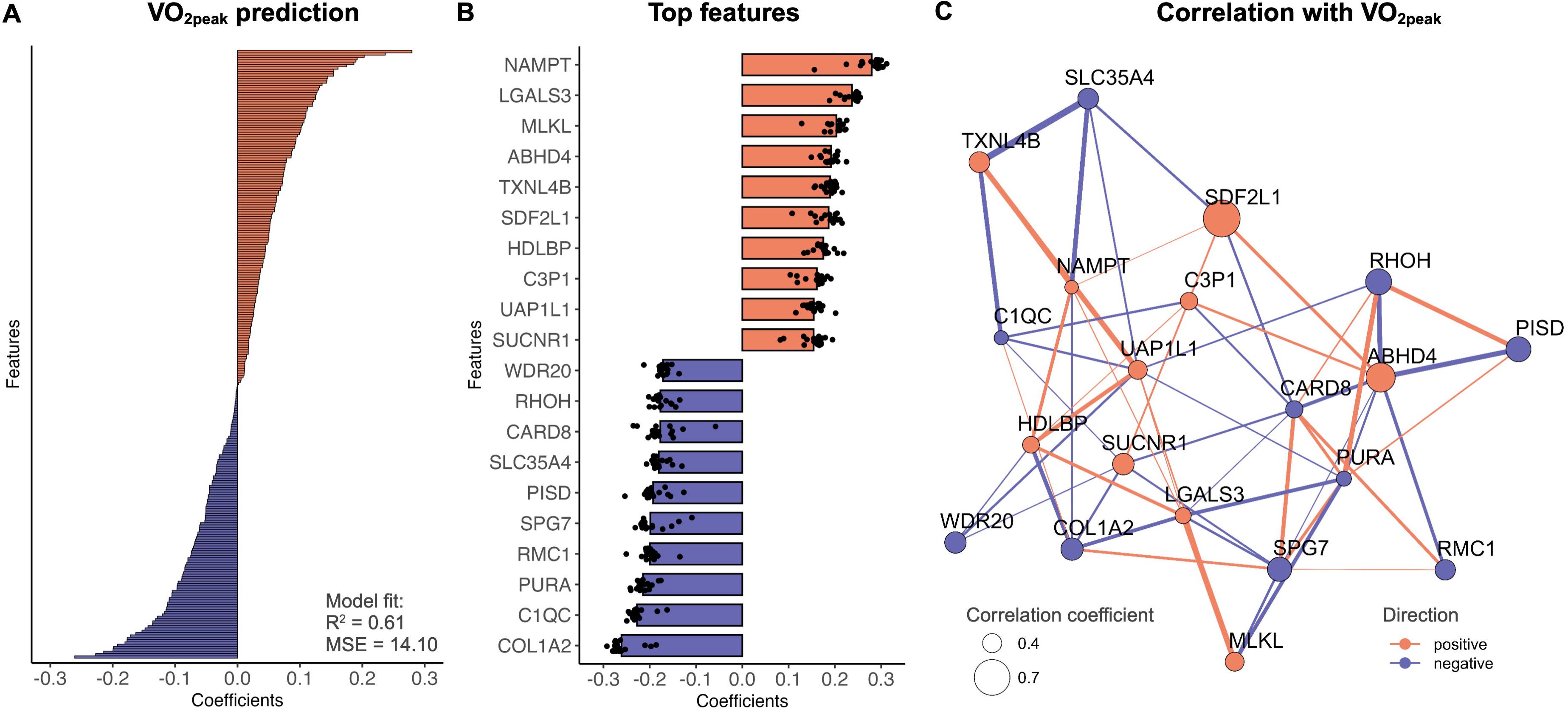
Identification of an immunoproteomic signature associated with cardiorespiratory fitness. (A) Ridge regression coefficients of features showing moderate to high correlation with VO2peak (r_S_ > 0.4 or < –0.4). (B) Ridge regression coefficients of the 20 features with highest predictive power for VO2peak. (C) Correlation network of the 20 features with highest predictive power for VO2peak. Dot sizes and colors represent the strength and direction of correlation with VO2peak. Connection width and colors represent the strength and direction of correlation between features. Connections are displayed for r_S_ > 0.3 or < –0.3.

We then focused our attention on the 20 features with the highest predictive power (Figure 6B). Interestingly, nicotinamide phosphoribosyltransferase (NAMPT), a key enzyme of nicotinamide adenine dinucleotide (NAD^+^) metabolism, demonstrated the highest positive impact on VO2peak prediction. NAMPT plays a crucial role in salvaging intracellular NAD^+^ and was previously shown to have a high association with exercise, as it forms a crucial component of energy metabolism in skeletal muscle,^49–51^ but also other target tissues like immune cells.^52–54^ Our results support this notion and suggest that repeated exposure to exercise, which results in greater cardiorespiratory fitness, equips immune cells with a higher metabolic capacity, thereby possibly linking to the immune effector functions that were previously identified in this work. Additionally, several studies have suggested a direct antiviral function of NAMPT in host defense.^55,56^ Similarly, we observed a positive impact on VO2peak prediction for succinate receptor 1 (SUCNR1), a G-protein coupled receptor that was previously shown to control exercise capacity and systemic glucose homeostasis in mice.^57,58^ There are several reports that the effects of exercise-secreted succinate on skeletal muscle tissue adaptions are dependent on paracrine signaling to non-myofibrillar cells such as macrophages, that express SUCNR1.^58,59^ SUCNR1 signaling is also critically involved in anti-inflammatory adaptions of adipose-tissue-resident macrophages,^60^ which directly links the secretion of succinate to anti-obesity effects and supports our findings that SUCNR1 is positively associated with VO2peak. Further proteins with positive impact on VO2peak prediction that support the notion of improved immunological health as a consequence of exercise training are stromal cell derived factor 2 like 1 (SDF2L1), a molecular component of the BiP chaperon cycle that prevents aggregation of misfolded proteins,^61,62^ and high density lipoprotein binding protein (HDLBP), which is suspected to prevent an over-accumulation of intracellular cholesterol.^63^ Besides features with positive impact on VO2peak prediction, our analysis also yielded several proteins with a negative impact (Figure 6B). Among these features phosphatidylserine decarboxylase (PISD) and caspase recruitment domain family, member 8 (CARD8) stood out due to their involvement in metabolism and inflammation. The negative impact of PISD, an enzyme involved in lipid droplet biogenesis,^64^ might be explained by reduced cellular fat deposition with increasing cardiorespiratory fitness. Thus, both HDLBP and PISD, despite showing contrary effects on VO2peak prediction, suggest adaptions in lipid metabolism of immune cells in response to exercise training. Additionally, CARD8, a pattern recognition receptor that mediates inflammasome activation and production of pro-inflammatory cytokines, also demonstrated a negative impact on VO2peak prediction. This suggest that besides metabolic adaptions, higher cardiorespiratory fitness might also be marked by anti-inflammatory adaptions, which confirms previous findings on the anti-inflammatory effects of exercise training.^12,65^ Finally, we generated a correlation network of features that showed high impact on VO2peak prediction and observed various positive and negative inter-feature correlations (Figure 6C).

Collectively our results suggest that the identified proteins associated with cardiorespiratory fitness reshape the phenotype of immune cells in response to exercise training. In taking an unbiased approach towards biological baseline characteristics of young healthy adults, we identified an immunoproteomic signature associated with cardiorespiratory fitness that is composed of 260 proteins, some of which are related to energy metabolism and anti-inflammatory adaptions (Figure 6C; Table S10). In a broader context these findings might serve as a molecular foundation for immunological health in the context of long-term training adaptions, thereby adding an immunological dimension to the well-described health benefits of exercise.^2^

## DISCUSSION

A better understanding of the molecular underpinnings of physical exercise in humans is needed to individualize exercise training recommendations and maximize their efficacy in mediating health benefits. While some human studies have addressed exercise responses and adaptions in skeletal muscle and blood plasma using state-of-the-art systems biology approaches,^6,9,66–68^ the impact of exercise on the immune system is less well understood. Since immune effector functions are directly involved in immunological defense, anticancer immunity, and anti-inflammatory adaptions, the immunological response to exercise is of high relevance for the prevention and treatment of numerous diseases. Here, we provide a comprehensive resource on how two different acute aerobic exercise stimuli rewire the proteomic makeup of PBMCs. We used a robust randomized crossover design, including two standardized baseline measurements, with a large sample size for proteomic analyses in human experimental interventional research. The findings of our study expand the literature by uncovering > 1000 proteins in PBMCs that change in response to acute exercise. Particularly, immune effector function and cell activation pathways are regulated in response to acute exercise, and higher intensity is needed to stimulate these changes. Finally, features derived from the PBMC proteome at baseline can be used to predict VO2peak, indicating a close association between immune cells and cardiorespiratory fitness on a proteomic level.

The acute exercise-induced mobilization of different immune cell subsets, in particular effector cells like NK cells, cytotoxic (CD8+) T cells and neutrophils,^11,12^ is well-investigated, but less is known on changes in their proteomic makeup and the resulting cell functions. Although a functional characterization of immune cell subsets is beyond the scope of this study, we identified comprehensive alterations in the immune cell proteome associated with cell function and activation pathways that match previous studies evaluating functional outcomes in different effector populations.^69–71^ The regulation of immune effector function and cell activation pathways indicate a (transient) state of immunomodulation following acute bouts of exercise, thereby possibly representing a mechanism of action underlying the benefits of exercise training and physical activity on cancer prevention and viral infections.

Interestingly, our results indicate that the observed changes in the proteomic makeup of PBMCs occur independent of exercise-induced alterations in PBMC composition. This is demonstrated by the fact that we observed far more alterations in protein abundance in response to HIIE compared to MICE, although the underlying PBMC shifts were identical between the two exercise conditions. Additionally, we identified several biological processes, that – when compared with immune cell mobilization – did not mirror the kinetic. For instance, we found a decreased regulation of “T cell receptor signaling pathway” 1h after HIIE (Table S9), but this timepoint was marked by an increased proportion of CD4+ T cells (Figure 1E). In contrast, “NK cell mediated immunity” was decreased (Table S9), which might be caused by the lower frequency of NK cells 1h after HIIE (Figure 1E). This shift in PBMC composition was neglected in previous investigations,^13^ but is a crucial component of exercise immunology that must be considered when interpreting alterations in immune cell transcripts, proteins, or effector functions over time. A further piece of evidence contradictory to a pure regulation by PBMC shifts arises from the fact that we found signature proteins specific to immune cell populations in our dataset that did not follow the corresponding kinetic of mobilization. By mining the human protein atlas^72,73^ for immune cell-specific proteins, we found an increased abundance of serpin family E member 1 (SERPINE1) – a protein with 25-fold higher expression in NK cells compared to the next highest cell type – 1h after HIIE (Table S5), although this timepoint was marked by a decreased proportion of NK cells (Figure 1E). Comparing our results to a previously published bulk RNA sequencing dataset that demonstrated large alterations in PBMC transcripts 2 minutes after exercise^13^ further demonstrates that our proteomics results are temporally in line with these transcriptomic alterations. In this context, our open resource offers the opportunity to mine the underlying dataset for specific proteins of interest. Our results can be used to inform new hypothesis-driven research in the field of exercise immunology and additionally allow detailed analyses on the consistency of immune cell protein levels between two standardized baseline measurements within the same human subjects as well as exercise-induced alterations and intensity-dependent differences.

Our experimental study results support the WHO recommendations on physical activity and sedentary behavior, which highlight the superior role of high exercise intensity for health promotion.^14^ From an immunological perspective, we found distinct responses of HIIE and MICE when matching the interventions for workload and duration and thus conclude that exercising at higher intensity is crucial to induce changes in the PBMC proteome. This might serve as a potential biological foundation for a recent comprehensive analysis revealing a superior effect of exercise intensity versus volume on longevity at a population-based level.^74^

Finally, to explore a potential association between two organ systems that are directly related to health and disease – the immune system and the cardiorespiratory system – we performed correlation and prediction analyses with VO2peak using all baseline data. While previous work has elucidated the molecular underpinnings of cardiorespiratory fitness by applying plasma proteomics and PBMC transcriptomics,^9,13,68^ a possible link between immune cell proteomics and cardiorespiratory fitness has not yet been explored. Interestingly, both, the correlation analysis as well as the prediction models, suggested strong associations with VO2peak. From all input features, particularly proteomics-derived features showed a strong positive or negative impact on VO2peak prediction. Highly predictive proteins such as NAMPT are crucial for cellular metabolism and we have recently shown that NAMPT gene expression and protein abundance of human PBMCs can be modulated by acute exercise.^53^ Overall, this suggests an interrelation between acute exercise stimuli, immunometabolic competence, and cardiorespiratory fitness and suggests a putative role of PBMCs as peripheral mirror for systemic health. Taken together, we find that PBMC proteome-derived features can predict VO2peak, thus open exciting research avenues on the molecular interconnection between immune cell proteostasis and cardiovascular fitness.

Of note, our study should be interpreted against the backdrop of some limitations. First, the temporal resolution is limited to 1h post exercise and many proteins that significantly changed in response to exercise did not return to baseline within this time frame. Future trials are needed to explore proteomic alterations at a higher temporal resolution using more follow-up measurement timepoints. Second, although we uncovered numerous protein changes and alterations in many functional pathways of PBMCs in response to HIIE, we cannot directly conclude on functional aspects of circulating immune cells. Additionally, as with every study using PBMCs as biomaterial, it is impossible to direct link protein changes to specific immune cell populations. Ultimately, it also remains questionable how exercise-induced alterations in PBMC composition affected our proteomic results. Compared to other studies in the field,^13^ we have addressed this caveat by quantifying alterations in PBMC composition and conclude that HIIE alters the proteome of immune cells much stronger than MICE despite identical immune cell mobilization patterns.

## CONCLUSION

In conclusion, we identified > 1000 exercise-induced alterations in the PBMC proteome and provide a valuable data resource for future research purposes. The identified changes were particularly related to immune cell effector function pathways, serving as a mechanistic link to the preventive and therapeutic impact of physical activity and regular exercise in various chronic diseases. In line with the WHO 2020 guidelines on physical activity and sedentary behavior, acute exercise at higher intensity elicited greater changes in the regulation of cell function and activation pathways, providing supportive biological evidence for the relevance of exercise intensity as an important factor when planning and structuring exercise training programs for health promotion. Finally, the associations between the PBMC proteome and VO2peak indicate systemic interrelations between the immune and cardiorespiratory system that can be modulated by physical exercise.

## SUPPLEMENTAL INFORMATION

Table S1. Overview of participant characteristics, related to Figure 1.

Table S2. Immune cell counts from flow cytometry analysis, related to Figure 1 and S1

Table S3. Detailed statistical results from analysis of exercise-induced changes in immune cell counts, related to Figure S1.

Table S4. Lymphocyte proportions, related to Figure 1.

Table S5. Detailed statistical results from analysis of exercise-induced changes in protein abundance of PBMCs, related to Figure 2 and 3.

Table S6. Detailed results of Gene ontology over-representation analysis, related to Figure 3.

Table S7. Membership values and fold changes from baseline of all proteins used for fuzzy c-means clustering, related to Figure 4.

Table S8. Detailed results of biological theme comparison, related to Figure 4.

Table S9. Detailed results of all Gene ontology gene set enrichment analyses, related to Figure 5, S3, S4, and S5.

Table S10. Detailed results of VO2peak prediction, related to Figure 6.

## Data Availability

All data produced in the present work are contained in the manuscript or available online.

https://www.ebi.ac.uk/pride

## ACKNOWLEDGEMENTS

We want to thank Lars Donath and Ludwig Rappelt for their help in conducting the trial. We thank the team of the Proteomics Core Facility of the DKFZ particularly Adrian Stoegbauer und Alina Ertl for sample preparation and LC-MS measurement. We also want to thank the German Sport University Cologne for supplying internal funds to A.J.M.. Figures were created with https://BioRender.com.

## AUTHOR CONTRIBUTIONS

Conceptualization, N.J., A.J.M., and P.Z.; Methodology, N.J., A.J.M., A.S., and P.Z.; Software, D.W., C.We., M.S., and S.C.; Formal Analysis, D.W., S.P., C.We., M.S., and S.C.; Investigation, A.J.M., S.P., A.S., M.S., and D.H.; Resources, C.Wa., C.A.O., D.H., and P.Z.; Writing – Original Draft, D.W., N.J., S.P., C.We., M.S., and S.C.; Writing – Review & Editing, A.J.M., S.P., A.S., C.We., A.L.H., M.S., S.C., A.G, C.Wa., C.A.O., D.H., and P.Z.; Visualization, D.W. and C.We.; Supervision, P.Z., A.G., and D.H., Project Administration, P.Z.; Funding Acquisition, A.J.M

## RESSOURCE AVAILABILITY

### Lead contact

Any requests for further pieces of information or resources should be directed to the Lead Contact Philipp Zimmer

### Data and code availability

Raw data files of all samples processed in the proteomics analysis are hosted on the PRoteomics IDEntifications Database (PRIDE; https://www.ebi.ac.uk/pride) under the following PRIDE-ID: PXD058573.

## EXPERIMENTAL MODEL AND STUDY PARTICIPANT DETAILS

### Participant recruitment and informed consent

Prior to enrollment of the first participants the study received ethical approval by the local ethics committee of the German Sport University Cologne, which works according to the World Medical Association’s Declaration of Helsinki. The study meets the National Institutes of Health definition of a clinical trial and was prospectively registered in the German Clinical Trials Register (DRKS00017686). Study eligibility was assessed for 28 healthy adults aged between 18 and 35. To ensure complication-free execution of the high-intensity interval exercise on the treadmill, participants required a weekly running volume of 2-5 hours and a body mass index < 30. Any previous medical history of muscle disorders, cardiac or kidney diseases as well as regular intake of medication or nutritional supplements were treated as exclusion criteria. For female participants, breast-feeding or an ongoing pregnancy were also treated as exclusion criteria. Of the 28 subjects assessed for eligibility, two were considered ineligible due to acute infections. The remaining 26 participants provided written informed consent and were enrolled in the study. After baseline testing two further participants dropped out due to orthopedic problems while running (Achilles injuries). For one participant, biomaterial did not suffice to run analyses, which resulted in a total of 23 participants. An overview of all participant characteristics is displayed in Table S1.

## METHOD DETAILS

### Study design

Participants enrolled in this randomized crossover study were scheduled for three visits to an exercise physiology laboratory of the German Sport University Cologne: Baseline testing, a HIIE session, and a MICE session. For each visit, participants were asked to arrive overnight-fasted and refrain from alcohol and caffein intake in the 24h prior. Water intake was permitted ad libitum. All visits were scheduled between 07:00 and 10:00 am to account for a potential circadian impact on performance and biological outcomes. The minimum timeframe between each of the three visits was 72 hours, to prevent potential carryover effects.

### Baseline testing

During baseline testing, written informed consent was obtained from participants and demographic, and anthropometric characteristics were recorded. Afterwards, participants underwent cardiopulmonary exercise testing.

#### Cardiopulmonary exercise test (CPET)

To standardize the exercise intensity between participants for the HIIE and MICE session, respectively, cardiorespiratory fitness was assessed as peak oxygen consumption (VO2peak) in an incremental CPET during baseline testing. The CPET was performed on a motorized treadmill (Woodway ELG 90, Weil am Rhein, Germany) that was set to 1 % incline for all sessions. The warm-up consisted of 5 min at 6-8 km h^-1^. Afterwards, participants began running at 8 km h^-1^. The speed of the treadmill was then increased by 1 km h^-1^ every 60 seconds until participants reached volitional exhaustion. During the test, heart rate was recorded continuously (Polar FS1C, Kempele, Finland), and rate of perceived exertion was recorded prior to each increase in intensity. Participants were verbally encouraged to continue running by the supervising researcher. After reaching volitional exertion, participants were given a 5 min break before taking up exercise again for a VO2peak verification test. For this test, the speed of the treadmill was set 1 km h^-1^ higher than what the participants had finished with. Just before the verification test, participants ran for 3 min at 8 km h^-1^. The speed was then increased to the target speed within 20 seconds and participants were instructed to run as long as possible. During the entire CPET participants wore a face mask that was connected to a spirometer (Cortex Metalyzer 3B, CORTEX Biophysik GmbH, Leipzig, Germany) to collect breathing gases breath-by-breath. The highest 15-second interval during the CPET was used to calculate VO2peak.

### Randomization

To prevent sequence effects arising from the order in which HIIE and MICE were conducted, participants were randomized into one of two exercise intervention sequences after baseline testing: HIIE-MICE or MICE-HIIE. Following the minimization procedure by Pocock and Simon,^75^ randomization was performed via concealed allocation (1:1) using the software Randomization in Treatment Arms (RITA; Evidat, LulJbeck, Germany). Age, BMI, and VO2peak were used as stratification factors.

### Exercise interventions

Exercise intensities for the HIIE and MICE session were calculated as percent of VO2peak for each participant to ensure that all participants exercised at the same intensity. The exercise protocols for HIIE and MICE were designed in an time– and workload-matched manner as previously described^15,16^ to isolate exercise intensity as the only differing variable between the two exercise conditions. Both exercise sessions were performed on the same treadmill that was also used for the CPET at baseline (Woodway ELG 90, Weil am Rhein, Germany). During MICE participants performed a warm-up for 10 min at a self-selected intensity, followed by a 5 min break. Participants then ran for 50 min at 70 % of their VO2peak. During HIIE, participants performed 7 min of warm-up and cool-down at 70 % VO2peak with six bouts of high-intensity running at 90 % VO2peak in between. Each high-intensity bout lasted 3 min, followed by 3 min of active recovery at 50 % VO2peak.

### Blood collection and sample preparation

Blood was drawn from a median antecubital vein in supine position at baseline, immediately after exercise, and 1h after exercise for the HIIE and MICE session, respectively. Each blood draw consisted of 24 mL of whole blood collected in EDTA tubes (Vacutainer, BD). After the last blood draw, peripheral blood mononuclear cells (PBMCs) were isolated via density gradient centrifugation. To achieve this, whole blood was first diluted with phosphate buffered saline (PBS) and then carefully layered on top of a lymphocyte separation medium (Cytiva Ficoll-Paque^TM^ PLUS, Fisher Scientific). After centrifugation for 30 min at room temperature and 800 g^-1^, the PBMC-containing interphase was collected, washed with PBS, and centrifuged again for 10 min at room temperature and 800 g^-1^. PBMCs were then resuspended in freezing medium (Recovery^TM^ cell culture freezing medium, Thermo Fisher Scientific) and stored at –80 °C before being transferred to a –150 °C freezer on the next day until further analysis.

### Flow cytometry

#### Sample preparation and data acquisition

Flow cytometry analysis was performed using a Cytek® Aurora full spectrum flow cytometer (Cytek Biosciences, California, USA). Cryopreserved PBMCs were gently thawed in a water bath at 37 °C with a mean recovery of 81.28 % viable cells assessed with the Zombie NIR™ Fixable Viability Kit (BioLegend, San Diego, CA, USA). After incubating 1 × 10^6^ PBMCs in 2.5 µg Fc block for 10 min at room temperature, cells were stained with anti-CD3 (BUV395, clone SK7), anti-CD4 (PerCP, clone SK3), anti-CD8 (BV750, clone SK1), anti-CD16 (PE-Cy7, clone 3G8), anti-CD25 (BUV805, clone M-A251), anti-CD56 (BUV563, clone NCAM16.2), anti-CD20 (APC, clone L27), and anti-CD19 (APC, clone SJ25C1) antibodies (all from BD Biosciences, NJ, USA). In brief, cells were incubated in the dark with a master mix containing Brilliant Stain buffer (BD Biosciences) and antibodies against surface antigens for 30 min at 4°C. After washing with FACS buffer, the BD Pharmingen™L Transcription Factor Buffer Set was used, and cells were fixed for 40 min at 4 °C in the dark. Thereafter, intracellular staining was done by incubating cells with an anti-Foxp3 antibody (PE, clone 259D/C7) for 45 min at 4 °C in the dark. After washing, cells were resuspended in FACS buffer and acquired on the flow cytometer within 2 hours after finishing the staining protocol.

#### Data processing

Gating was performed using FlowJo™ 10.10.0 (Figure 1C). B cells were phenotyped as CD3^-^CD56^-^CD19^+^CD20^+^, Natural Killer T (NKT) cells as CD3^+^CD56^+^, Natural Killer (NK) cells either as CD56^bright^CD16^-^ (NK^bright^) or CD56^dim^CD16^+^ (NK^dim^), and regulatory T cells (T_regs_) as CD4^+^CD25^+^Foxp3^+^. The person analyzing the samples was blinded to the participants’ group allocation. Analysis of total blood cell counts was performed from EDTA blood using a hematology analyzer (SYSMEX XP-300, Norderstedt, Germany). The lymphocyte count was then used to calculate the absolute number of peripherally circulating lymphocyte subsets according to the cell proportions derived by flow cytometry.

### LC-MS/MS-based untargeted proteomics

Sample were processed and measured in a block randomized order^76^ to prevent any technical bias that might occur during sample preparation or LC-MS measurement.

#### Sample preparation

Isolated PBMCs were lysed in a RIPA buffer containing 10 mM sodium fluoride, 1 mM sodium orthovanadate, cOmplete^™^ Protease Inhibitor Cocktail (Merck KGaA), PhosSTOP^™^ (Merck KGaA), 250 U mL^-1^ benzonase, and 10 U mL^-1^ DNase I. Samples were incubated on ice for 1h and then centrifuged at 4 °C and 13,000 g^-1^ for 15 min. Protein concentration was determined in the supernatant with a BCA assay. An amount of 10 µg of protein per sample was digested (Trypsin) using an AssayMAP Bravo liquid handling system (Agilent technologies) running the autoSP3 protocol.^77^ After sample preparation the remaining peptides were vacuum dried and stored at –20 °C until LC-MS/MS analysis.

#### MS method Orbitrap Exploris 480

The dried peptide sample was reconstituted (97.4 % Water, 2.5 % Hexafluoro-2-propanol and 0.1 % trifluoroacetic acid (TFA)) and 10 % of the sample were used. The LC-MS/MS analysis was carried out on an Ultimate 3000 UPLC system (Thermo Fisher Scientific) directly connected to an Orbitrap Exploris 480 mass spectrometer for a total of 120 min. Peptides were online desalted on a trapping cartridge (Acclaim PepMap300 C18, 5 µm, 300 Å wide pore; Thermo Fisher Scientific) for 3 min using 30 µl/min flow of 0.1 % TFA in water. The analytical multistep gradient (300 nL/min) was performed using a nanoEase MZ Peptide analytical column (300Å, 1.7 µm, 75 µm x 200 mm, Waters) using solvent A (0.1% formic acid in water) and solvent B (0.1% formic acid in acetonitrile). For 102 min the concentration of B was linearly ramped from 4 % to 30 %, followed by a quick ramp to 78%, after two min the concentration of B was lowered to 2 % and a 10 min equilibration step appended. Eluting peptides were analyzed in the mass spectrometer using data independent acquisition (DIA) mode. A full scan at 120 k resolution (380-1400 m/z, 300 % AGC target, 45 ms maxIT) was followed 47 DIA windows. The DIA acquisition covered a mass range of 400-1000 m/z using windows of a variable width with 1 m/z overlap, an AGC target of 1000 % with a maxIT set to 54 ms and recorded at a resolution of 30 k. Each sample was followed by a wash run (40 min) to avoid carry-over between samples. Instrument performance and suitability was monitored by regular (approx. one per 48 hours) injections of a standard sample and an in-house shiny application over the whole timeline of the experiment.

#### Data analysis

Analysis of DIA RAW files was performed with Spectronaut (Biognosys, version 19.1.240724.62635)^78^ in directDIA+ (deep) library-free mode. Default settings were applied with the following adaptions. Within DIA Analysis under Identification the Precursor PEP Cutoff was set to 0.01, the Protein Qvalue Cutoff (Run) set to 0.01 and the Protein PEP Cutoff set to 0.01. In Quantification the Proteotypicity Filter was set to Only Protein Group Specific, the Protein LFQ Method was set to MaxLFQ and the quantification window was set to Not Synchronized (SN 17). The data was searched against the human proteome from Uniprot (human reference database with one protein sequence per gene, containing 20,597 unique entries from ninth of February 2024) and the contaminants FASTA from MaxQuant (246 unique entries from twenty-second of December 2022).

#### Data processing

Before further analysis, the obtained dataset was checked for proteins that were identified more than once. For these duplicate results, the event with the highest number of identified precursors across all samples was kept and all other events were deleted from the dataset. The data was then filtered for proteins that were quantified in ≥ 70 % of the samples in at least one exercise condition and measurement timepoint (i.e., HIIE/MICE baseline, post exercise, 1h post exercise). Subsequently, we imputed the data separated by exercise condition and measurement timepoint using the missForest package.^79^ Ultimately, proteins were annotated to match the gene names provided in the org.Hs.eg.db package for subsequent Gene Ontology (GO) analysis. Translation between gene names and Entrez gene identifiers was accomplished with the bitr function from the ClusterProfiler package.^46,80^

## QUANTIFICATION AND STATISTICAL ANALYSIS

Samples from a total of 23 participants were available for statistical analyses. For one participant there was no sample from 1h after MICE due to difficulties during PBMC isolation. Statistical analysis and visualization were performed in R. If not otherwise noted, data wrangling was achieved using the dplyr^81^ and tidyr^82^ package and subsequently visualized with ggplot2^83^ and ggpubr^84^.

### Unsupervised immune cell clustering using self-organizing maps

Flow cytometry data of each sample was cleaned using the FlowAI plugin (v3.2.3) in FlowJo™ 10.10.0. The remaining events were gated as described above and live cells were downsampled to 3,000 events per sample using the DownSample plugin (v3.3.1). Subsequently, downsampled events were concatenated to obtain an overall dataset containing all exercise conditions (HIIE, MICE) and measurement timepoints (baseline, post exercise, 1h post exercise). This dataset was then used to perform unsupervised immune cell clustering using self-organizing maps (SOM) with the FlowSOM plugin (v4.1.0). The resulting 6 clusters were identified as CD4+ T cells, CD8+ T cells, NKT cells, CD56^dim^ cells, CD56^bright^ cells and B cells in the build-in Cluster Explorer in FlowJo™ 10.10.0. The overall dataset was visualized using Uniform Manifold Approximation and Projection (UMAP) via the UMAP plugin (v4.1.1) and FlowSOM clusters were superimposed via color-coding. This overall, color-coded immune cell clusters were used as a template map for subsequent clustering per exercise condition and measurement timepoint. To achieve this, the downsampled events were concatenated for baseline, post exercise and 1h post exercise in HIIE and MICE, respectively. Ultimately, FlowSOM clustering and UMAP were performed on each of these concatenated dataframes by applying them on the previously generated map.

### Exercise-induced mobilization of immune cells

Exercise-induced alterations in immune cell counts were analyzed by applying linear mixed models to the flow cytometry results. Measurement timepoint and exercise condition were implemented as fixed effects and participant ID as random effect using the lmer function from the lme4 package.^85^ Results of the linear mixed models were then analyzed for time and time × condition interaction effects via analyses of variance (ANOVAs) with the built-in ANOVA function from R stats. In case of significant results, pairwise comparisons of measurement timepoints and/or exercise conditions were performed by applying the emmeans function from the emmeans package. P-values were Bonferroni-corrected for multiple testing.

### Measures of variability

All measures of variability were calculated with unimputed data to avoid potential bias arising from imputation. Inter-individual variability was calculated as coefficient of variation (CVs) for each protein across all participants separated by exercise condition (HIIE, MICE), and measurement timepoint (baseline, post exercise, 1h post exercise). CVs were calculated as the ratio of the standard deviation σ to the mean *µ*. Intra-individual variability was assessed by comparing the baseline values of the two intervention days. Relative differences between day 1 and day 2 were calculated (in percent) for each protein separated by study participant. Proteomic variability was quantified for each protein by calculating (i) the mean CV across all participants in HIIE and MICE at baseline, (ii) the mean CV across all participants in HIIE and MICE post exercise and 1h post exercise, and (iii) the mean difference between the two baselines across all participants. Proteomic variability was also calculated separated by exercise conditions and measurement timepoints (see Figure S2A and S2B).

### Principal component analysis

Principal component analysis (PCA) was carried out using the built-in prcomp function from R stats. All samples were plotted with the fviz_pca_ind function from the factoextra package. Exercise condition, measurement timepoint, intervention day, and sex were used as metadata to color-code PCA results. PCAs were also computed on datasets separated by exercise condition or measurement timepoint to visually assess the impact of these variables on each other (see Figure S2C-E).

### Linear mixed models to identify proteins altered by HIIE and/or MICE

To identify proteins altered by HIIE and/or MICE, a linear mixed model was fitted on the log_2_-transformed, normalized, and imputed protein intensities via the limma R package.^86^ Intra-individual correlation was estimated via the duplicateCorrelation function. The model included the exercise condition (HIIE, MICE), the measurement timepoint (baseline, post exercise, 1h post exercise), and the interaction between both as fixed factors. A moderated *t* statistic^87^ was obtained for each contrast of interest via the eBayes function with estimated variance trend and robustification. The resulting p-values for each contrast were adjusted with the Benjamini-Hochberg procedure^88^ to control the false discovery rate and significance was declared at the adjusted 5 % two-sided level. The model was subsequently extended to include sex and all two-way interactions.

### Gene ontology (GO) over-representation analysis

Time effects of the statistical analysis with limma^86^ were used to map proteins that were significantly altered by HIIE and MICE to GO terms, respectively. GO over-representation analysis was performed with the ClusterProfiler package.^46,80^ For HIIE and MICE, significantly altered proteins were compared with the entire dataset of quantified proteins applying Benjamini-Hochberg correction of p-values with a p-value cutoff of 0.05 and a q-value cutoff of 0.2.

### Fuzzy c-means clustering

Fuzzy c-means clustering was performed with the Mfuzz package.^89,90^ Data was standardized using the standardise function and the optimal number of clusters was determined by calculating the minimum centroid distance for a range of cluster numbers using the Dmin function. The optimal fuzzifier was identified with the mestamiate function.

### Biological theme comparison

Biological theme comparison was carried out using the compareCluster function from the ClusterProfiler package.^46,80^ Entrez gene identifiers of the proteins contained in the identified clusters were used as input with the function command set to “enrichGO”. Benjamini-Hochberg correction was applied to p-values with a cutoff of 0.05 and minimum gene set size was set to 10. The results were simplified via the simplify function using a cutoff of 0.7 and visualized separated by ontology with the cnetplot function from the enrichplot package.^91^

### Gene ontology (GO) gene set enrichment analysis

Gene set enrichment analysis was performed using the gseGO function from the ClusterProfiler package.^46,80^ Entrez gene identifiers and fold changes from baseline of the proteins contained in the identified clusters were used as input with the minimum gene set size set to 10. In case fold changes were only positive or negative, the “scoreType” command was set to “pos” or “neg”, respectively. P-values were corrected using the Benjamini-Hochberg procedure with a p-value cutoff of 0.05. The underlying proteins mapping to each significant GO term were identified using the select function from the AnnotationDbi package. Shared and unique GO terms across the identified clusters were visualized with the UpSetR package.^92^

### VO2peak prediction and correlation network analysis

#### Preselection of features

To identify features with high association to VO2peak, we conducted a preselection in Python (v.3.9).^93^ The features were standardized using z transformation and included the average of mass spectrometry-based proteomics data and flow cytometry-based immunophenotyping data at baseline of intervention day 1 and 2 as well as sex, height, weight and BMI. VO2peak was scaled to body weight. Data from 2 participants were excluded from the analysis due to incomplete feature sets. Pairwise Spearman’s rank correlations between all features and VO2peak were calculated (Table S10) and features with a correlation coefficient of > 0.4 or < –0.4 were included in the subsequent analysis. From a total of 6,063 initial features, 260 remained after this selection.

#### VO2peak prediction modeling

We ran LASSO^94,95^ and ridge regression^96^ as well as a random forest^97^ as a non-linear, tree-based approach. A leave-one-out (LOO) cross-validation was performed in Python (v.3.9) to assess the predictive performance of these methods based on the 260 features. To optimize the hyperparameters for each model by grid search, a second inner cross validation was performed. For each training set, we selected the model that had the lowest test error. The predicted output value resulted from the cross validation iteration, where the corresponding output data point and its associated features were not included in the training set. These predicted values were used to calculate the mean squared error (MSE) and the r squared R^2^. Ridge regression outperformed the other models. All features with coefficients from the ridge regression are listed in the supplements (Table S10).

#### Correlation network analysis

The 20 features with the largest absolute mean value from the ridge regression were selected to create a weighted, undirected network using Spearman’s rank correlations. The network was visualized in R (v.4.4.1) with the packages Hmisc (v.5.2.1) and igraph (v.2.1.1.).

## FIGURE LEGENDS

**Figure S1.**
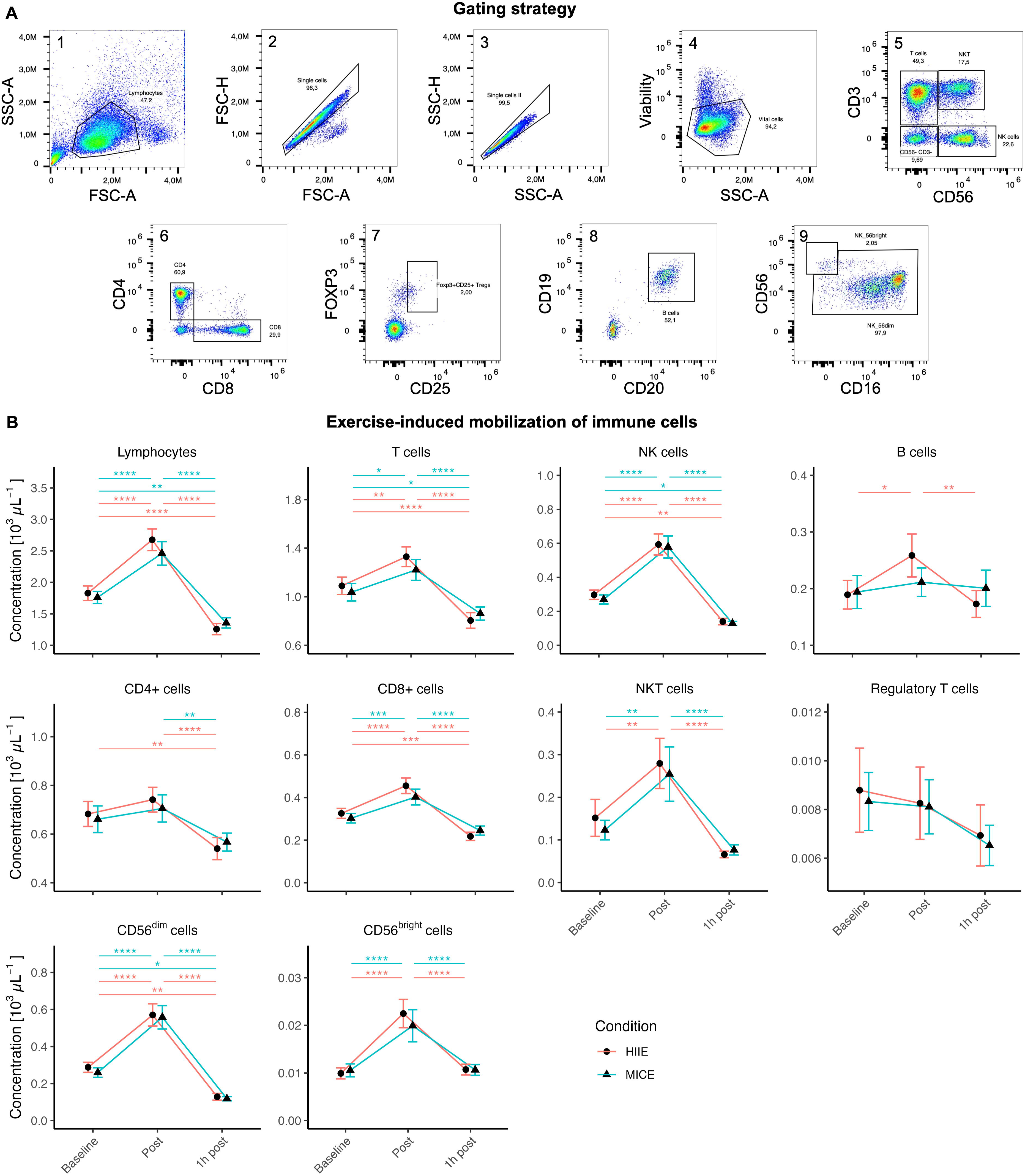
Immune cell mobilization is independent of exercise intensity, related to Figure 1 and Table S2, S3, and S4. (A) Gating strategy applied to PBMCs. Gated immune cell populations included vital lymphocytes, T cells, CD4+ T cells, CD8+ T cells, natural killer (NK) T cells, regulatory T cells, NK cells, CD56^dim^ cells, CD56^bright^ cells, and B cells. (B) Mobilization kinetics of total lymphocytes and lymphocyte subsets in response to high-intensity interval exercise (HIIE) and moderate-intensity continuous exercise (MICE). Linear mixed models with subsequent analyses of variance and Bonferroni-corrected pairwise comparisons were applied. Data are represented as mean ± SEM (n = 22). **** p < 0.0001, *** p < 0.001, ** p < 0.01, * p < 0.05.

**Figure S2.**
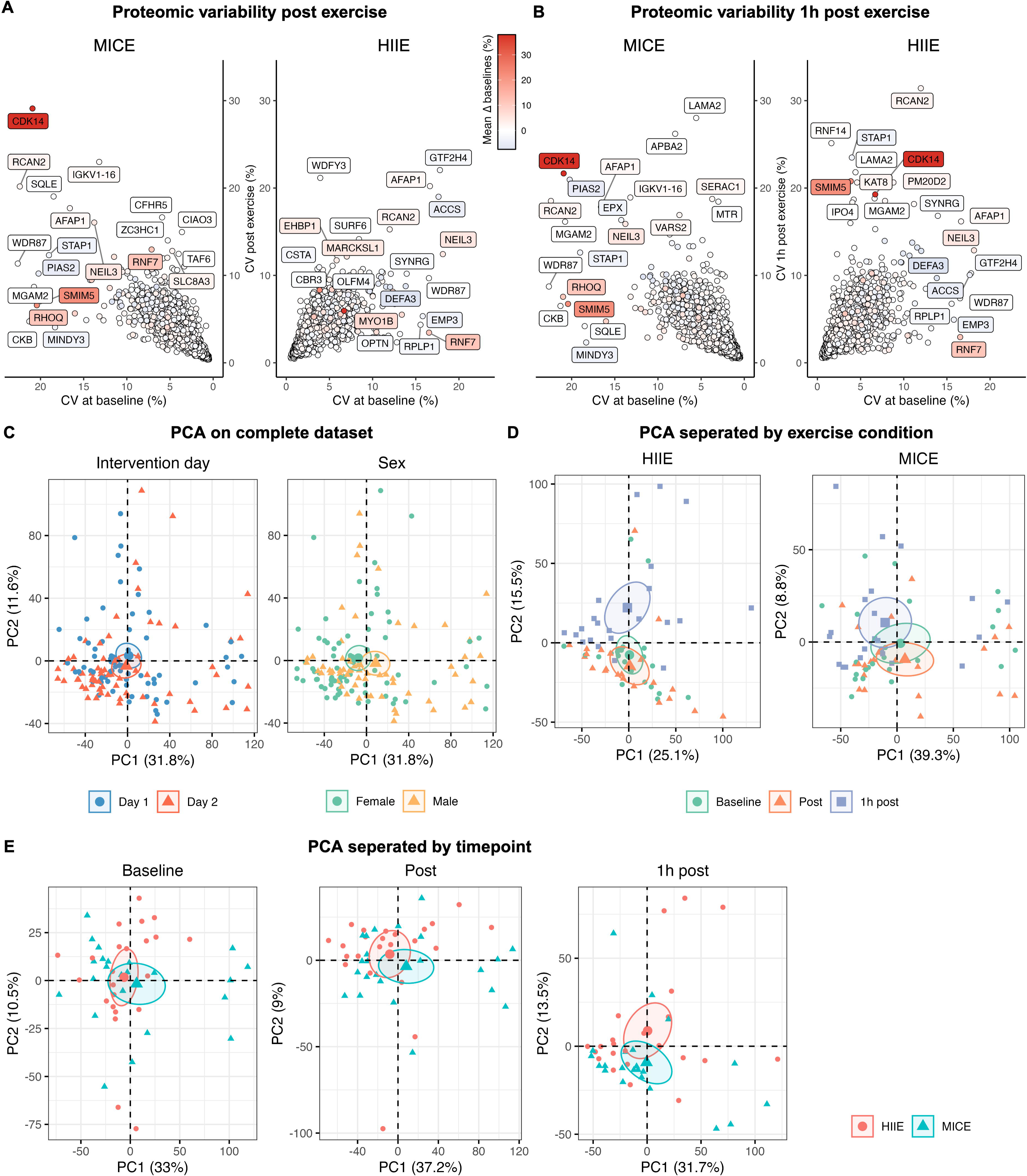
Proteomic variability and principal component analyses (PCA) separated by exercise condition and measurement timepoint, related to Figure 2. (A) Proteomic variability immediately after HIIE and MICE. (B) Proteomic variability 1h after HIIE and MICE. (C) PCA on complete dataset to evaluate the impact of intervention day and sex. (D) PCA separated by exercise condition to evaluate the impact of measurement timepoint. (E) PCA separated by measurement timepoint to evaluate the impact of exercise condition.

**Figure S3.**
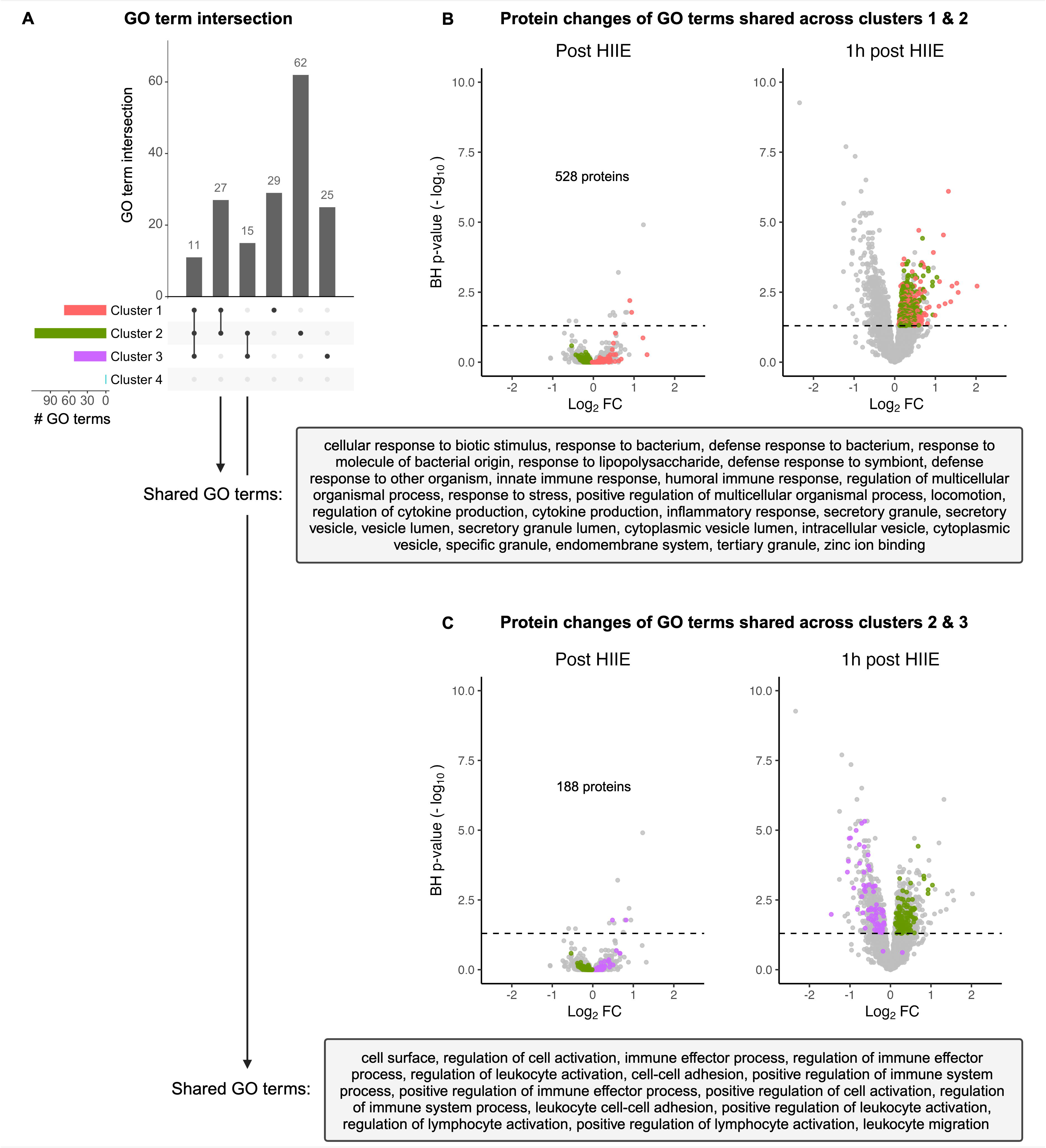
GO terms overlap between clusters 1 & 2 and 2 & 3, respectively, related to Figure 5 and Table S9. (A) Overview of shared and unique GO terms across all clusters. (B) GO terms shared between clusters 1 and 2. (C) GO terms shared between clusters 2 and 3.

**Figure S4.**
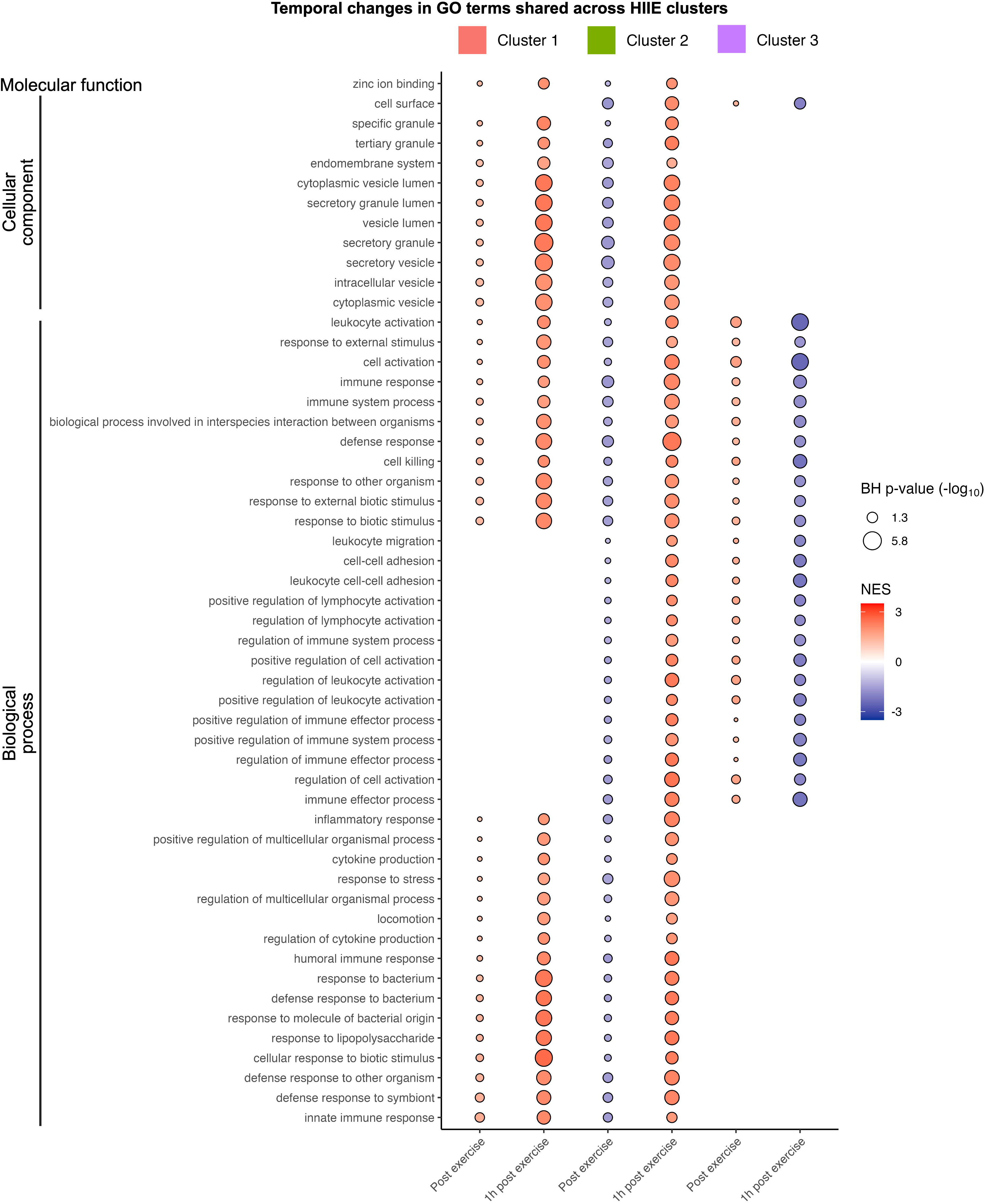
Temporal regulation of GO terms shared across protein clusters 1 – 3, related to Figure 5 and Table S9.

**Figure S5.**
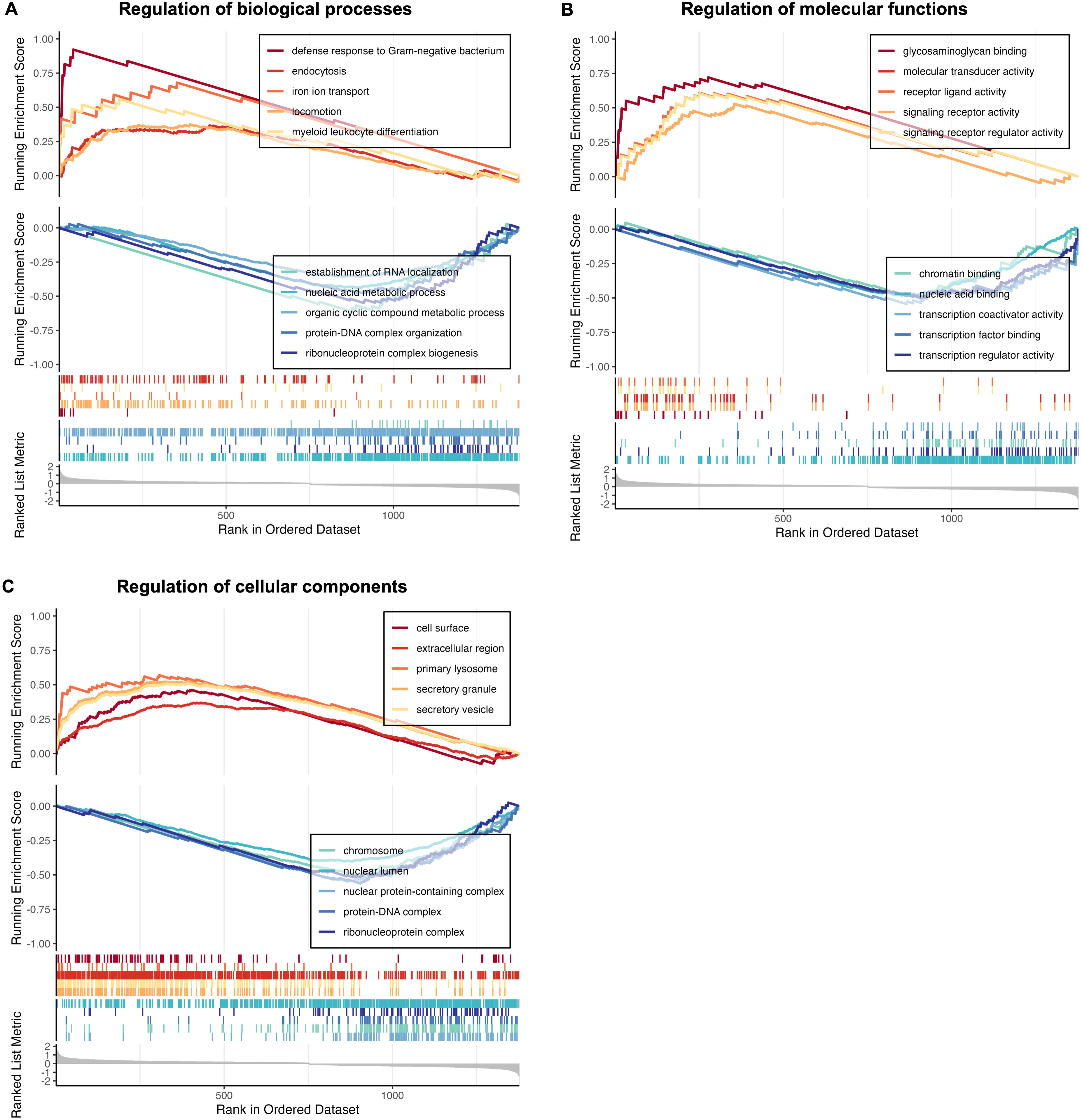
Global gene set enrichment analysis suggests regulation of immune cell effector function and transcriptional regulation in response to HIIE, related to Figure 5, S3, and S4 and Table S9. (A) Highest and lowest enrichment scores for regulation of biological processes. (B) Highest and lowest enrichment scores for regulation of molecular functions. (C) Highest and lowest enrichment scores for regulation of cellular components.

